# Enhanced machine learning and hybrid ensemble approaches for coronary heart disease prediction

**DOI:** 10.1101/2025.07.02.25330708

**Authors:** Maurice Wanyonyi, Zakayo Ndiku Morris, Faith Mueni Musyoka, Dominic Makaa Kitavi

## Abstract

**Introduction:** Coronary heart disease (CHD) remains the leading cause of mortality worldwide, disproportionately affecting low- and middle-income countries where diagnostic resources are limited. Traditional statistical models often fail to deliver adequate predictive accuracy in complex, high-dimensional, and imbalanced health datasets.

**Objective:** To develop and evaluate enhanced machine learning and hybrid ensemble models for the prediction of coronary heart disease, with a focus on improving diagnostic performance, interpretability, and applicability in resource-constrained settings.

**Methods:** We utilized a nationally representative dataset of 253,680 individuals from the Behavioral Risk Factor Surveillance System. Preprocessing included normalization and balancing via the Synthetic Minority Oversampling Technique (SMOTE). Baseline models—Decision Trees, Random Forests, Gradient Boosting, and Support Vector Machines—were compared against improved versions: Adaptive Noise–Resistant Decision Tree (ADNRT), Hybrid Imbalanced Random Forest (HIRF), Pruned Gradient Boosting Machine (PGBM), and Enhanced Support Vector Machine (ESVM). Ensemble approaches (stacking, boosting, bagging, Bayesian model averaging and majority voting) were implemented and evaluated using accuracy, sensitivity, specificity, and area under the curve (AUC). Calibration and learning curves were also analyzed.

**Results:** Enhanced models consistently outperformed their baseline counterparts. PGBM achieved the highest sensitivity (90.8%), while HIRF demonstrated the best overall calibration and balance (AUC = 0.937; sensitivity = 88.4%; specificity = 82.9%). The stacking ensemble emerged as the best-performing model with an accuracy of 87.2%, sensitivity of 89.6%, specificity of 84.7%, and AUC of 0.94. Calibration and learning curve analyses confirmed strong generalizability and low overfitting across ensemble models.

**Conclusion:** Hybrid ensemble machine learning models significantly outperform traditional classifiers in CHD prediction, offering high accuracy, robustness, and interpretability. These models present a scalable framework for implementing AI-driven diagnostic tools in low–resource environments, potentially transforming early detection and prevention of coronary heart disease.

## Introduction

The most widespread type of cardiovascular disease (CVD), coronary heart disease (CHD), has been the primary cause of death globally since it kills more than 7 million people each year and accounts for around 85 per cent of all deaths associated with cardiovascular issues [1], [43]. Though much progress has been made in the field of cardiovascular care, the incidence of CHD is increasing, especially in low- and middle-income countries (LMICs), where there is still limited access to services that can help prevent and diagnose CHD at an early stage and treat it adequately [7], [30].

Demands for effective, scalable, and legible tools that enable early prediction and intervention are more emergent now than ever. In sub-Saharan Africa (SSA), CHD is becoming more prevalent among younger, economically productive groups, further increasing the public health and economic consequences [27], [45]. CHD has reached epidemic proportions in Kenya, leading to an increase in the number of hospital admissions and heart failure, which have been worsened by risk factors, including hypertension, obesity, diabetes, and HIV-related complications [14], [32].

Early CHD mortality risk prediction is of immense importance in reducing mortality and improving patient outcomes, especially in situations with resource constraints.

Logistic regression and Cox proportional hazards models are among the statistical models traditionally used to identify primary cardiovascular disease (CHD) risk factors and predict outcomes [24], [21]. These models are appreciated because they are interpretable and straightforward; however, in high-dimensional health data situations, they tend to misrepresent the complex and nonlinear relationships that exist therein. Over the past few years, extensive research has been conducted on machine learning (ML) methods due to their ability to learn from data, detect previously unknown patterns, and achieve improved predictive performance [16], [25]. The random forest, support vector machines, gradient boosting, and multi-layer perceptron algorithms have shown promising outcomes in CHD prediction [2], [8], [41].

Nevertheless, there are a few limitations to machine learning (ML) models. Most of them face problems of overfitting, lack of transparency, and inadequate generalization when used in different populations, particularly in low- and middle-income countries (LMICs), where the quality of data and its completeness may not be improved [4, 6]. In addition, although a wide variety of research has been conducted to develop ML-based diagnostic technologies, there is still a distinct research gap in models aimed at the predictive progression of CHD, an indispensable consideration when it comes to the application of preventative and long-term management of disease [10], [23]. Moreover, ML models have been commonly assessed in isolation, without being fully integrated and compared with classical statistical methods; their contextual applicability and interpretability in clinical decision-making are inevitably constrained.

To bridge this gap in methodology, the ensemble learning approach has emerged as an influential strategy that integrates the strength of single models in order to reduce their deficiencies. Different ensemble methods, including bagging, boosting, and stacking, have been shown to significantly outperform predictive models across diverse clinical settings [22], [46], [39]. These techniques attempt to fuse predictions of many base learners, where the base learners are usually a combination of interpretable models, like logistic regression, and strong learners, like random forest and XGBoost, to deliver robustness, better performance, and generalization. Specifically, stacked generalization has proven to achieve better performance in predicting CHD because it learns to assign the best possible weights to the predictions of base models, minimizing both bias and variance at the same time [34], [8].

Current literature underscores the advantages of ensemble approaches in cardiovascular risk modeling. For instance, [22] proposed an improved ensemble method based on CART models, achieving accuracy scores of over 93% on benchmark datasets. [39] developed a stacked ensemble model that outperformed standalone classifiers, reporting an accuracy of 92.34%. Likewise, [8] validated ensemble methods that utilize feature selection and hyperparameter tuning to achieve robust classification performance. Meanwhile, Shorewala 2021 found that ensemble models outperformed traditional base classifiers by an average of 1.96% in accuracy. These findings collectively suggest that integrating multiple predictive paradigms can significantly enhance diagnostic power and mitigate the limitations of individual classifiers.

Nevertheless, the majority of these ensemble strategies have been tested on high-income datasets and often lack contextual relevance to LMICs such as Kenya. Most models are designed with little consideration for resource constraints, data sparsity, and infrastructural limitations commonly encountered in LMIC healthcare systems [26], [28]. Furthermore, limited research has explored how ensemble models perform when classical statistical models–typically favored for their interpretability—are integrated with complex ML models in a structured hybrid framework. This integration could potentially balance the trade-off between prediction accuracy and interpretability, a crucial requirement in healthcare settings where clinicians must trust and understand model outputs.

Recent advances in heart disease prediction have incorporated explainable AI (XAI), hybrid learning frameworks, and generalizable deep models. For instance, CardioRiskNet [35] combines attention mechanisms, active learning, and XAI to deliver highly accurate and interpretable risk predictions yet focuses primarily on deep architectures and does not benchmark against traditional or ensemble-based learners. Similarly, the HXAI-ML framework [36] integrates advanced resampling strategies and SHAP/LIME/PIA explanations, achieving superior results on benchmark datasets but without clustering or ensemble fusion layers. [38] demonstrate that boosting algorithms like XGBoost and CatBoost perform well in CHD prediction, identifying key clinical features with support from conformal classifiers, though their work does not explore hybrid ensemble construction or subgroup analysis. Meanwhile, [31] propose a transparent prediction interface tailored for clinicians using XAI and user-centric design but with limited performance benchmarking across multiple learner types.

In this study, we seek to address existing methodological and contextual gaps in CHD prediction by developing a hybrid modeling approach that integrates diverse machine learning algorithms into robust ensemble frameworks. Specifically, our study pursues two primary objectives: (1) to evaluate the predictive performance of individual machine learning models using a large, population-based dataset; and (2) to develop ensemble models that combine these algorithms to enhance both prediction accuracy and interpretability.

To ensure robustness and relevance, the study employs a comprehensive methodological framework, including preprocessing (handling missing data, class imbalance, and outliers), diagnostic statistics, feature selection, and hyperparameter tuning through grid search. The individual models explored include decision trees, random forests, support vector machines, and gradient boosting machines. These are evaluated using standard metrics such as accuracy, precision, recall, F1-score, and area under the receiver operating characteristic curve (AUC–ROC). This multi-layered strategy aims not only to improve prediction performance but also to provide interpretable insights that can be acted upon in real–world clinical settings.

In contrast to recent studies–such as CardioRiskNet [35], HXAI-ML [36], and other explainable AI frameworks [38], [31], our work addresses three key gaps. (1) The development of enhanced machine learning variants—including Adaptive Noise Resistant Decision Trees (ANRDT) and Hybrid Imbalanced Random Forests (HIRF)—specifically tailored for CHD prediction in resource-constrained environments; (2) A comprehensive benchmarking of ensemble strategies, including advanced techniques such as Bayesian Model Averaging, to assess predictive performance and generalization across models; and (3) The integration of unsupervised clustering analysis to identify latent CHD risk subgroups, offering both predictive enhancement and clinically meaningful subgroup interpretation. Collectively, these contributions advance the methodological landscape of CHD prediction by improving accuracy, interpretability, and contextual relevance—especially within low- and middle-income country (LMIC) settings such as Kenya.

Accordingly, this work provides a structured comparison between individual machine learning models, their enhanced variants, and hybrid ensemble frameworks. Unlike prior studies that either focus solely on standalone models or lack contextual relevance to LMICs, this study emphasizes methodological rigor and practical utility. By benchmarking these models on a standardized dataset and evaluation framework, the research contributes actionable insights for developing scalable, interpretable, and accurate CHD prediction systems to support early detection and improved clinical decision-making in resource-limited settings.

## Materials and methods

### Data Source

dary coronary heart disease dataset sourced from the IEEE Dataport repository, a well–established platform for sharing high-quality datasets in health research, including epidemiological and cardiovascular disease studies [47]. The selected dataset (the Behavioral Risk Factor Surveillance System (BRFSS) dataset) was collected by the U.S. Centers for Disease Control and Prevention (CDC) and includes responses from 253,680 individuals, with 22 variables encompassing demographic, behavioral and lifestyle, clinical information, and healthcare access & general well-being. Out of the total records, 229,787 do not indicate CHD, while 23,893 are classified as CHD-positive based on self-reports.

The decision to use BRFSS dataset is driven by both its comprehensive nature and the relevance of its variables to CHD prediction in diverse populations. Despite being U.S.–based, BRFSS captures risk factors (e.g., smoking, cholesterol levels, blood pressure, diabetes) that are also prevalent in LMICs. Given the limited availability of large-scale, population-level CHD datasets in LMICs like Kenya, BRFSS serves as a valuable data source for model training. The model can later be fine-tuned and validated using smaller, country-specific datasets from LMICs to ensure adaptability and accuracy in local settings.

### Preprocessing

Missing values in the data was addressed using appropriate imputation techniques, such as multiple imputation or k–nearest neighbors (KNN), depending on the nature of the missing data pattern. The methods used to detect outliers included the interquartile range (IQR) and Mahalanobis distance. The presence of noise was eliminated using noise reduction techniques such as smoothing filters, feature correlation analysis, and variance thresholding. Lastly, normalization and standardization were employed to guarantee uniform scaling of features and enhance the model training performance.

### Handling Class Imbalance

In the original data, the CHD–positive cases class was extremely low in relative values to the total sample (less than 10 per cent of the total sample). This is why Synthetic

Minority Oversampling Tecnique (SMOTE) was employed. SMOTE creates synthetic samples of the minority class (CHD positive) by interpolation among existing minority observations in the instance (feature) space. In contrast to random oversampling, SMOTE stabilizes the risk of overfitting because it adds variability to the synthetic samples, thus resulting in a more balanced and generalizable decision boundary for the classifiers [5], [44]. Subsequently, SMOTE applied to the dataset returned a balanced dataset, which was used to develop a model since the classifier became more sensitive to CHD cases. At the same time, its specificity remained unaltered. SMOTE is a popular method used to address class imbalance in machine learning datasets by generating synthetic examples of the minority class. The basic mathematical equation behind SMOTE is as follows:

Let:

- *x*_*i*_ be a minority class sample (feature vector),
- *x*_nn_ be one of its *k*-nearest neighbors (also from the minority class),
- *λ* be a random number in the range [0, 1], Then a synthetic sample *x*_new_ is generated as:

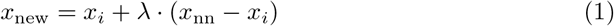

### Sample Size Determination

The original number of observations in the present research was 253,680. Following the balance between the classes using SMOTE, the size of the whole sample was significantly expanded to N = 459,574. This larger dataset presented a great computational challenge as it had to be balanced with a better representation. When trained on this complete dataset, machine learning algorithms required too many iterations to contend with and usually caused the system to crash or become unresponsive due to insufficient memory. To mitigate this, the Yamane formula was used to calculate the representative subsample, since it is a simplification of calculating the sample size of a given population and the level of precision that is desired [3], [40]. The formula is expressed as:

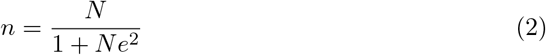

Where:

- *n* is the calculated sample size,
- *N* is the population size,
- *e* is the margin of error (level of precision). For this study:

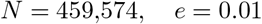

Substituting into the formula:

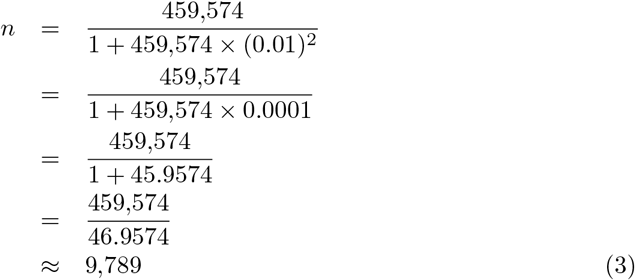

Therefore, the ultimate sample size chosen to train the machine learning models was about n = 9,789 observations. In order to make sure that this sample was representative enough of the structure and diversity of the whole dataset, especially in terms of class imbalance and essential demographic or clinical sub-populations, the stratified sampling method was used. Stratified sampling involves dividing the populations into uniform subunits or strata (e.g., by classes or labels, age intervals, or gender), and each of these strata is sampled proportionately [37], [17]. The method proves particularly useful within healthcare-driven data sets where minorities (i.e., disease outcomes) are poorly represented [15]. Stratified sampling increases representativeness and makes all significant subpopulations well-represented in the sample. It is primarily suggested to enhance the results of training machine learning models to eliminate bias and variance in unbalanced datasets [19].

### Base Model Selection

Several base machine learning models were considered in this study and include Decision Trees, Random Forests, Gradient Boosting Machines, and Support Vector Machines. The reason for selecting these models is their performance with complex and high-dimensional data, as well as their popularity among predictive modelling problems. Nevertheless, there are inherent limitations within each of these models that may contravene its performance, as noted in recent research. To mitigate these difficulties, better versions of the models have been put forward to make them more robust and more predictive.

**Decision Trees** are simple yet effective decision models that divide the data by performing feature thresholds and dividing the processed data into zones, each of which is associated with a specific value of the target. They are highly interpretable but also increase the risk of overfitting and are very sensitive to noise [6]. To enhance noise immunity and overfitting, this study utilized an Adaptive Noise-Resistant Decision Tree (ANRDT), which is an improved form of decision tree. In contrast to traditional trees, which apply hard threshold splits, ANRDT also applies probabilistic (soft) splits via a sigmoid function so that smooth decisions on the boundaries of the decisions can be made. By doing so, the model will be less reliant on low-level input data variances. Moreover, model complexity and generalization are regulated with pruning and regularization techniques, which help ANRDT to be more stable and tolerant of noise in real-world data. The formula for the improved decision tree is as follows:

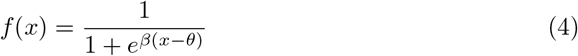

Where *f* (*x*) is the predicted probability, *β* is the steepness of the decision boundary, *x* is the predictor value, and *θ* is the split threshold.

**Random Forests** increase the stability and performance of a model by averaging out multiple predictions made by different decision trees. Nevertheless, they can have problems with skewed data and cannot give precise estimations of feature importance [11]. This study proposed a Hybrid Imbalanced Random Forest (HIRF) to solve the issues of class imbalance and interpretability associated with the standard Random Forests. HIRF improves on the performance by setting the SMOTE to equalize the underrepresented classes and make the models fair. Also, the SHapley Additive exPlanations (SHAP) is employed to assign meaningful scores to individual tree based on their contributions to the prediction and thus to enhance the interpretability of the feature importance. The weighted ensemble method, therefore, focuses on trees that extract important patterns in the data and produce predictions that are both more accurate and explainable.The HIRF model is given by

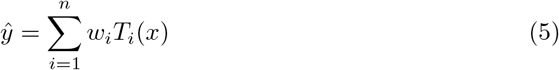

Where *ŷ* is the predicted target, *n* is the number of trees, *T*_*i*_(*x*) is the prediction from the *i*-th tree, and *w*_*i*_ is the weight assigned to each tree based on SHAP values.

**Gradient Boosting Machines:** iteratively construct decision trees, refining predictions by minimizing errors and focusing on residuals. While highly effective, GBMs are susceptible to overfitting and sensitive to outliers [20]. To mitigate these issues, this study implemented a Pruned Gradient Boosting Machine (PGBM), which integrates early stopping and Huber loss to enhance model robustness and generalization. The Huber loss function is defined as:

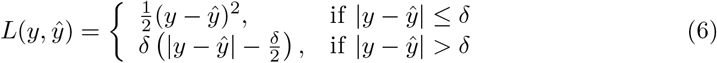

In this equation, *δ* is the threshold for identifying outliers, *y* is the true value, and *ŷ* is the predicted value.

Early Stopping monitors validation performance and stops training when further iterations no longer improve the model, preventing overfitting. Huber Loss Function combines Mean Squared Error (MSE) and Mean Absolute Error (MAE) to reduce the influence of extreme outliers while maintaining smooth gradient updates.

**Support Vector Machines(SVM):** are powerful supervised learning classifiers that construct a maximum-margin hyperplane to optimally separate data points of different classes. Despite their effectiveness, standard SVMs are often criticized for being difficult to interpret and for requiring meticulous hyperparameter tuning [4].

To address these limitations, this study introduced an Enhanced Support Vector Machine (ESVM) framework. The ESVM improves interpretability by incorporating SHapley Additive exPlanation (SHAP) values, which attribute the contribution of each feature to individual predictions. Additionally, the model employs Bayesian optimization to automatically tune hyperparameters, thereby improving efficiency and predictive performance.

The standard SVM seeks to find the optimal hyperplane by solving the following convex optimization problem:

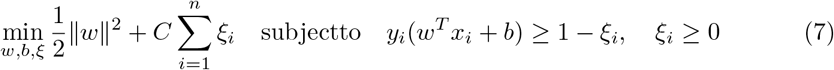

To enable non-linear classification, the ESVM utilizes the Radial Basis Function (RBF) kernel, which maps input features into a higher-dimensional space. The RBF kernel is defined as:

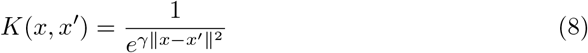

Where:

- *K*(*x, x*′) is the kernel function measuring similarity between input vectors *x* and *x*′,
- ∥*x* − *x*′∥^2^ is the squared Euclidean distance,
- *γ* is a hyperparameter controlling the kernel’s smoothness.

The dual optimization problem, often used for kernel-based SVMs, is formulated as:

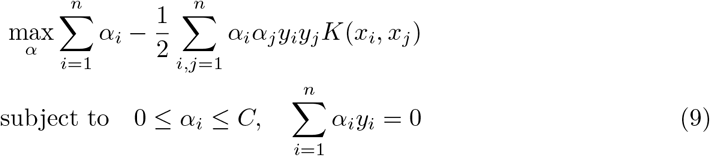

The resulting decision function used to classify new instances is given by:

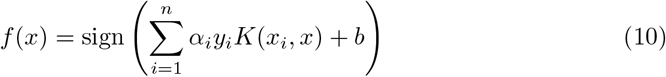

To enhance model transparency, the ESVM uses SHAP values to decompose the prediction:

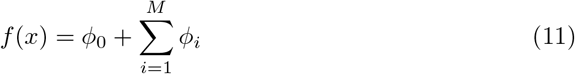

Where:

- *f* (*x*) is the predicted output,
- *ϕ*_0_ is the base value (expected prediction),
- *ϕ*_*i*_ is the SHAP value for feature *i*,
- *M* is the total number of input features.

This combination of SHAP-based interpretability and Bayesian hyperparameter tuning makes ESVM a robust and transparent model suitable for high-stakes domains such as healthcare prediction.

### Data Split and Model Training

In order to facilitate the adequate development and assessment of the model, the BRFSS dataset has been split into three subsets: 70% as training, 15% as validation and 15% as a test set. Such stratified split was able to create effective models without the danger of overfitting and guarantee the user-friendly applicability of the findings [13]. The base models were fitted on the training set, and the validation set was utilized to help fine–tune the hyperparameters and to help tune the model performance. The test set was not used in training and validation but instead was used as an independent test set to test the predictive power of the final model. Further improvement of the reliability was achieved by using a 5-fold cross-validation of the training phase. Here, the training data was split into five equal phases or “folds”. Each time there is an iteration, one fold should be kept as a temporary validation set, and the rest of the folds are utilized to train the model. This process is repeated five times, with each fold used as the validation set once. Cross–validation not only supports robust model tuning but also helps mitigate overfitting by ensuring that the model performs consistently across different subsets of the data. Model training aimed to identify the classifier that best predicts CHD outcomes based on the selected predictor variables while minimizing the loss function on the test set. Figure 1 shows the machine learning framework.

**Fig 1.**
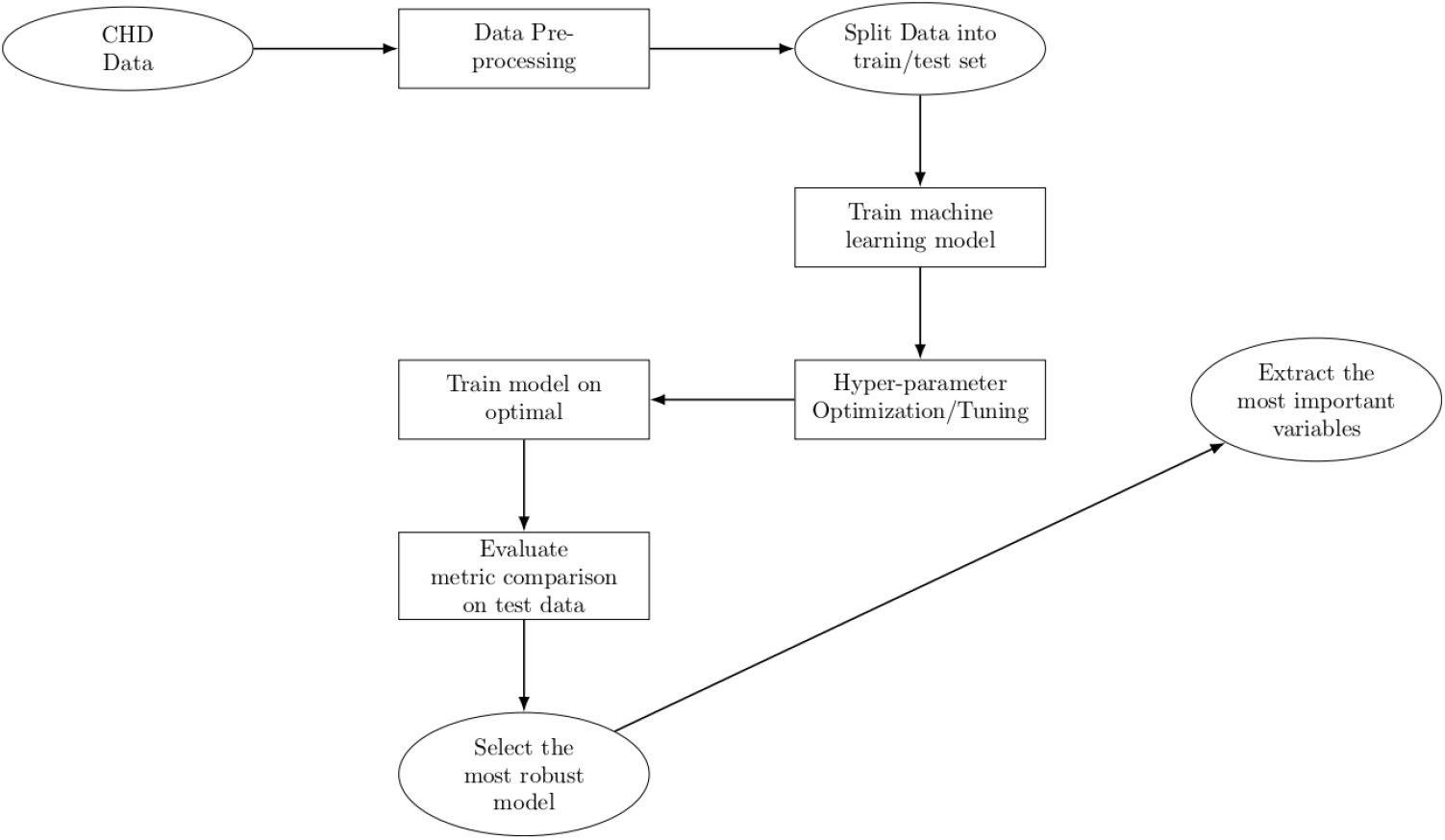
Machine Learning framework

### Clustering Analysis

To explore hidden subpopulations among individuals affected by coronary heart disease (CHD), an unsupervised clustering approach was applied independently across four domains: demographics, clinical characteristics, behavioral and lifestyle patterns, and healthcare access, combined with general well-being. A fifth integrative clustering was then performed using all variables to capture cross-domain profiles. The K–Means algorithm was selected for its efficiency and suitability for structured epidemiological data [12], partitioning the dataset into *K* clusters by minimizing the sum of squared distances between data points and their respective centroids, as described by the objective function:

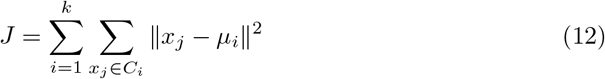

where *µ*_*i*_ denotes the centroid of cluster *i*, and *C*_*i*_ represents the set of data points assigned to that cluster. prior to clustering, all continuous features were standardized using Z-score normalization, while categorical variables were appropriately encoded to ensure balanced influence across the algorithm. Domain-specific clustering enabled focused analysis of key determinants: demographic attributes (e.g., age, sex, education, income), clinical indicators (e.g., blood pressure, cholesterol, diabetes), behavioral risk factors (e.g., smoking, diet, alcohol use), and access/well-being measures (e.g., insurance, general health, mental and physical health days). Optimal cluster numbers were identified using silhouette scores. To evaluate associations with CHD outcomes, Chi-square tests, and multivariable logistic regressions were conducted, with cluster labels as predictors. This multi-level strategy revealed nuanced patient subgroups, enhancing the understanding of how combined social, clinical, and behavioral factors shape CHD risk and guiding more precise cardiovascular prevention strategies.

### Ensemble Learning

To enhance the predictive performance of the ensemble framework, this study integrated multiple ensemble learning techniques: Boosting, Bagging, Stacking, Majority Voting, and Bayesian Model Averaging (BMA). Each technique was implemented and compared to determine the one that achieves the highest predictive accuracy and reliability for CHD.

**Boosting:** is an ensemble learning technique that aims to convert a set of weak learners into a strong learner through iterative refinement. Unlike bagging, where models are trained independently and in parallel, boosting trains models sequentially. Each new model is trained to correct the errors made by the previous ensemble of learners by giving more focus (weight) to the misclassified observations. Formally, given a training set

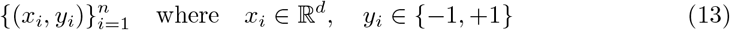

for binary classification, boosting builds an additive model of the form:

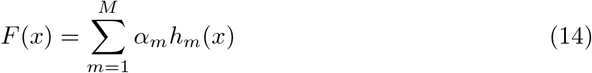

where:

- *h*_*m*_(*x*) is a weak learner (e.g., a shallow decision tree) at iteration *m*,
- *α*_*m*_ is the weight assigned to learner *h*_*m*_,
- *F* (*x*) is the final strong classifier after *M* iterations.

**AdaBoost:** In AdaBoost (Adaptive Boosting), the weights of misclassified samples are increased so that the next learner focuses more on difficult cases. The model minimizes the exponential loss:

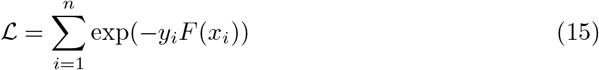

At each step:

1. The weak learner *h*_*m*_(*x*) is trained to minimize the weighted classification error:

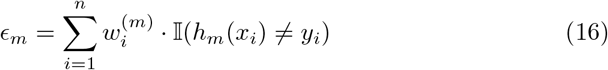
2. The model weight *α*_*m*_ is computed as:

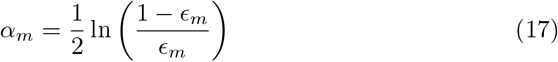
3. The sample weights are updated:

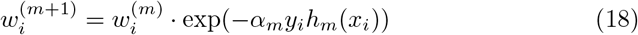

and then normalized to sum to 1.

**Gradient Boosting Machines:** Gradient Boosting generalizes boosting by optimizing a differentiable loss function ℒ (*y, F* (*x*)) using gradient descent. At each iteration, a new learner *h*_*m*_(*x*) is fitted to the negative gradient of the loss with respect to the current model prediction:

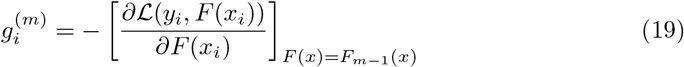

Then *h*_*m*_(*x*) is trained to predict 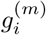 and the model is updated as:

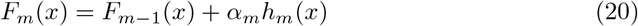

where *α*_*m*_ is the learning rate (step size). Popular implementations of boosting algorithms include XGBoost, LightGBM, and CatBoost, which introduce improvements in speed, scalability, regularization, and support for categorical features. These optimizations make gradient boosting one of the most powerful techniques in modern machine learning.

**Bagging (Bootstrap Aggregating):** is an ensemble learning technique that aims to reduce model variance and prevent overfitting by training multiple models on different bootstrap samples (random samples with replacement) of the original dataset. Each individual model, often referred to as a *base learner*, is trained independently, and their predictions are aggregated to produce the final output. For regression tasks, the final prediction is the average of the individual predictions:

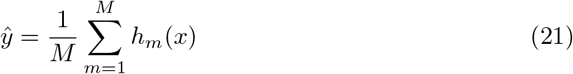

where *h*_*m*_(*x*) is the prediction from the *m*-th model and *M* is the total number of models. For classification tasks, majority voting is typically used:

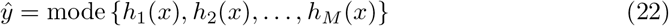

A widely–used bagging-based algorithm is **Random Forest**, which constructs an ensemble of decision trees. In addition to training on bootstrap samples, Random Forest introduces further randomness by selecting a random subset of features at each split in the tree-building process. This decorrelates the individual trees and further improves ensemble diversity, stability, and predictive accuracy.

**Stacking:** is a hierarchical ensemble learning technique that combines the predictions of multiple base models through a meta-model (also known as a *level-1 model*). Unlike bagging and boosting, which use homogeneous models and direct aggregation (e.g., averaging or voting), stacking allows heterogeneous base learners and learns how to best combine their predictions. In the first stage, *M* base models

*h*_1_(*x*), *h*_2_(*x*), …, *h*_*M*_ (*x*) are trained on the training data. The predictions of these models on a hold-out validation set are collected to form a new dataset:

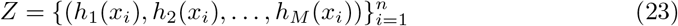

A meta-model *H*(*Z*) is then trained on this new dataset *Z*, learning to combine the outputs of the base models. The final prediction is:

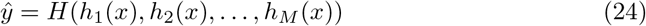

Stacking can be used for both classification and regression. In classification, the meta-model is often a logistic regression or support vector machine; in regression, it could be a linear regressor or gradient boosting machine.

**Majority Voting and Averaging:** are simpler ensemble strategies where no meta-model is involved. For classification, the final prediction is made based on the majority class predicted by the base models:

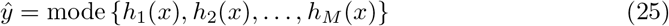

For regression, the predictions are averaged:

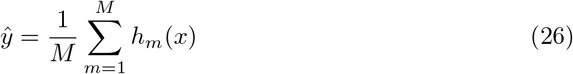

While majority voting and averaging improve robustness through consensus, stacking often yields better performance by learning complex interactions among model predictions.

**Bayesian Model Averaging (BMA):** is an ensemble learning technique that incorporates model uncertainty by averaging over multiple models, each weighted by its posterior probability given the data. Rather than selecting a single “best” model, BMA considers the contribution of all candidate models in making predictions, thus improving robustness and accuracy.

The final prediction is computed as a weighted sum of the individual model predictions:

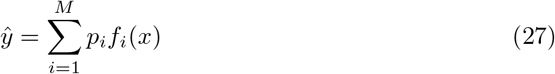

where:

- *f*_*i*_(*x*) is the prediction from the *i*-th base model,
- *p*_*i*_ = *P* (ℳ_*i*_ | *D*) is the posterior probability of model ℳ_*i*_ given the observed data *D*,
- *M* is the total number of models,
- *ŷ* is the final ensemble prediction.

The posterior probabilities satisfy the normalization constraint:

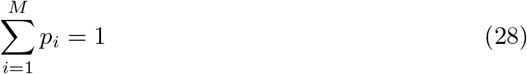

The posterior probability *p*_*i*_ for each model ℳ_*i*_ can be computed using Bayes’ theorem:

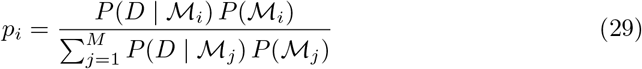

where *P* (*D* | ℳ_*i*_) is the marginal likelihood (model evidence) and *P* (ℳ_*i*_) is the prior probability of model _*i*_. BMA combines the strengths of multiple models while explicitly accounting for the uncertainty in model selection. This leads to more calibrated predictions and often outperforms model selection approaches that rely on a single model, especially when model evidence is comparable across candidates.

### Evaluation Metrics

The performance of the ensemble methods was compared with the classical statistical models and the individual machine learning models in terms of sensitivity, specificity, accuracy, F1– score, and AUC–ROC. These measures are developed based on the confusion matrix, which is a primary instrument in assessing the classification model performance. It gives a table of actual and predicted classifications, which offers an in-depth analysis of how the model works. Table 1 represents the confusion matrix that corresponds to the classification of CHD cases where the rows indicate the true classes of the CHD cases and the columns indicate the predicted classes of the CHD cases. The matrix is an overview of how well the model can differentiate between true negatives (non–CHD patients correctly predicted), true positives (CHD patients correctly predicted), false positives (non–CHD patients incorrectly classified as CHD) and false negatives (CHD patients not correctly predicted by the model).

**Table 1.** Confusion matrix structure.

|  | Predicted Positive | Predicted Negative |
| --- | --- | --- |
| Actual Positive | True Positive (TP) | False Negative (FN) |
| Actual Negative | False Positive (FP) | True Negative (TN) |

Table notes: This table outlines the standard structure of a binary classification confusion matrix, indicating the relationships between actual and predicted outcomes.

Sensitivity (true positive rate) measures the proportion of actual positive cases correctly identified by the model. A sensitivity of 1 signifies perfect identification of all true positives, while a value of 0 indicates that the model failed to identify any true positives.

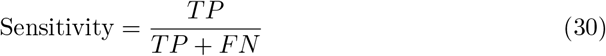

where *TP* = True Positives, *FN* = False Negatives.

Accuracy measures the proportion of correct predictions (both true positives and true negatives) among all predictions made. An accuracy of 1 indicates that the model made correct predictions for every case, while an accuracy of 0 means that every prediction was incorrect.

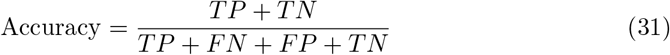

where *TN* = True Negatives, *FP* = False Positives.

Specificity (true negative rate) measures the proportion of actual negative cases correctly identified by the model. A specificity of 1 indicates perfect identification of all true negatives, while a value of 0 means the model failed to identify any true negatives.

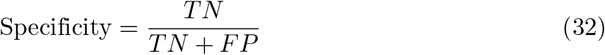

where *TP* = True Positives, *FN* = False Negatives.

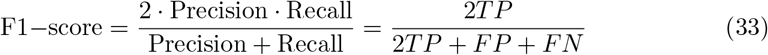

AUC-ROC (Area Under the Receiver Operating Characteristic curve) evaluates the model’s ability to discriminate between positive and negative cases. An AUC of 1 signifies a perfect model with optimal discrimination. A value of 0 means the model is completely ineffective and is classifying oppositely.

### Software and Analytical Tools

All data cleaning, preprocessing, modeling, and visualization were conducted using **Python version 3.13**. The analytical workflow integrated a suite of specialized libraries to ensure efficiency, accuracy, and reproducibility.

**Data manipulation and preprocessing** were performed using pandas and numpy, while `scikit-learn` was used for model building, evaluation, and implementation of resampling techniques such as Synthetic Minority Oversampling Technique (SMOTE).

**Ensemble and tree-based models** such as Random Forest, Gradient Boosting, and Stacking were implemented via `scikit-learn`, with enhanced versions (e.g., Pruned GBM and Noise-Resistant Trees) incorporating customized Python classes and hyperparameter tuning using `GridSearchCV` and `RandomizedSearchCV.`

**Data visualization** and exploratory analysis were conducted using `matplotlib, seaborn,` and `plotly,` with interactive visualizations used for exploratory cluster analysis and survival curve interpretation. Clustering and dimensionality reduction employed `scikit-learn’s KMeans,` and silhouette analysis functions.

All scripts were version-controlled using Git, and `Jupyter Notebooks` were used for iterative experimentation. Reproducibility was further ensured through fixed random seeds, isolated virtual environments (via `venv` or `conda`), and consistent preprocessing pipelines.

### Ethical Consideration

Ethical approval was deemed unnecessary for this study despite involving human subjects because the data used was secondary, and there was no physical contact with human subjects during data collection/retrieval. The data can be availed to anyone upon subscription at IEEE*Dataport* (Heart Disease Dataset)

## Results

### Data Balancing and Class Distribution

Initially, there was a significant class imbalance, with only 9.42% of individuals classified as having experienced heart disease or a heart attack (Yes = 1). The vast majority (90.58%) were classified as not having experienced such events (No = 0). This reflects a common issue in medical and epidemiological datasets, where the condition of interest occurs in a relatively small population subset. Such imbalance poses a significant challenge for machine learning models, particularly those relying on classification, as they tend to be biased toward predicting the majority class. This can lead to high overall accuracy but poor sensitivity (recall) for the minority class, which, in this context, is the class of most significant clinical interest. The SMOTE technique was applied to generate synthetic examples of the minority class. As seen in Table 2, the result was a perfectly balanced distribution of 50.0% for each class, thereby ensuring that the classifier receives equal representation of both outcomes during training. This balanced distribution improves the model’s ability to learn minority class patterns and predict positive cases of CHD more effectively.

**Table 2.** Class distribution of *Heart Disease or Attack* before and after SMOTE.

| Heart Disease or Attack | Before SMOTE (%) | After SMOTE (%) |
| --- | --- | --- |
| No (0) | 90.58 | 50.00 |
| Yes (1) | 9.42 | 50.00 |

Table notes: This table presents the percentage distribution of the target classes before and after applying the SMOTE technique to balance the dataset.

The bar plots generated before and after applying SMOTE provide a clear visual illustration of the balancing process in the dataset as shown in Figures 2 and 3 respectively. Before SMOTE, the bar representing class 0 (individuals without heart disease or heart attack) was dramatically taller than that of class 1 (those with the condition), visually emphasizing the significant class imbalance. This aligns with the earlier numerical summary, which indicated that nearly 91% of the cases were negative. In contrast, the bar plot after applying SMOTE shows both classes represented by bars of equal height, reflecting a perfectly balanced distribution of 50% for each class. This visual transformation confirms the successful oversampling of the minority class and highlights the importance of addressing such imbalance in predictive modelling.

**Fig 2.**
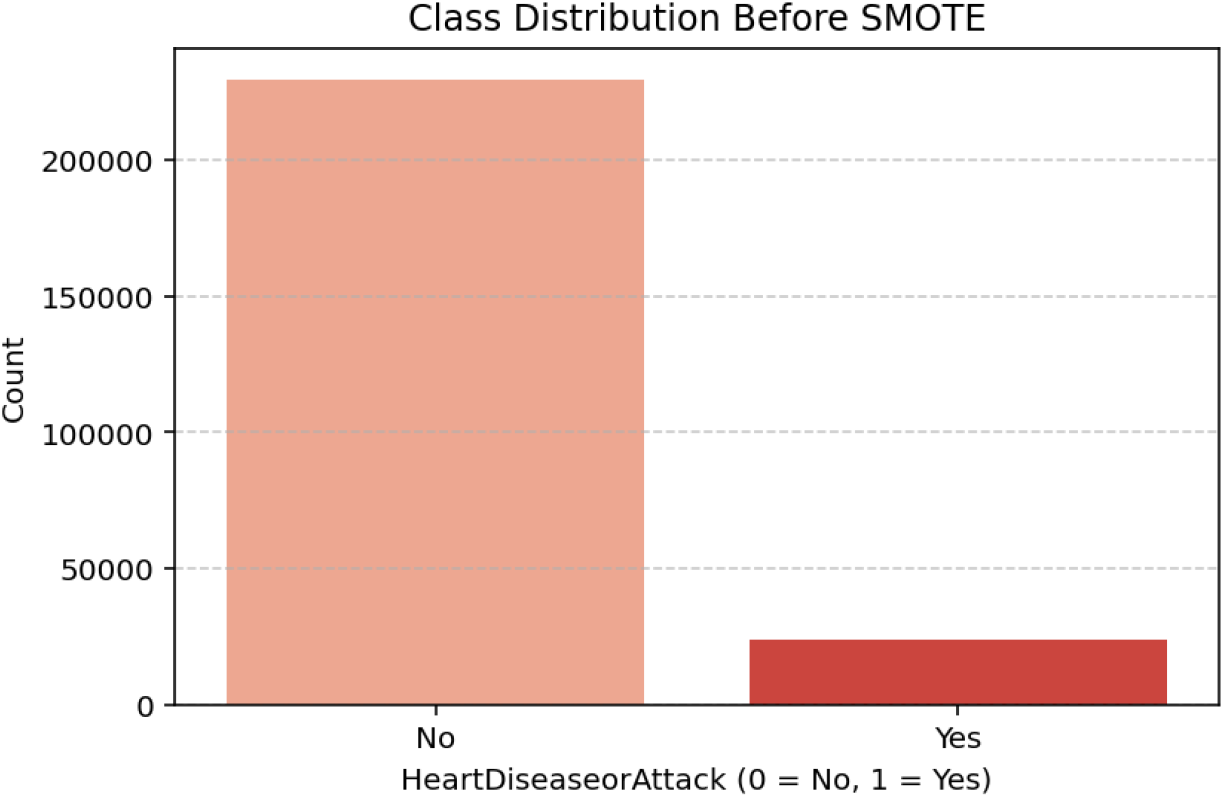
Class Distribution Before SMOTE.

**Fig 3.**
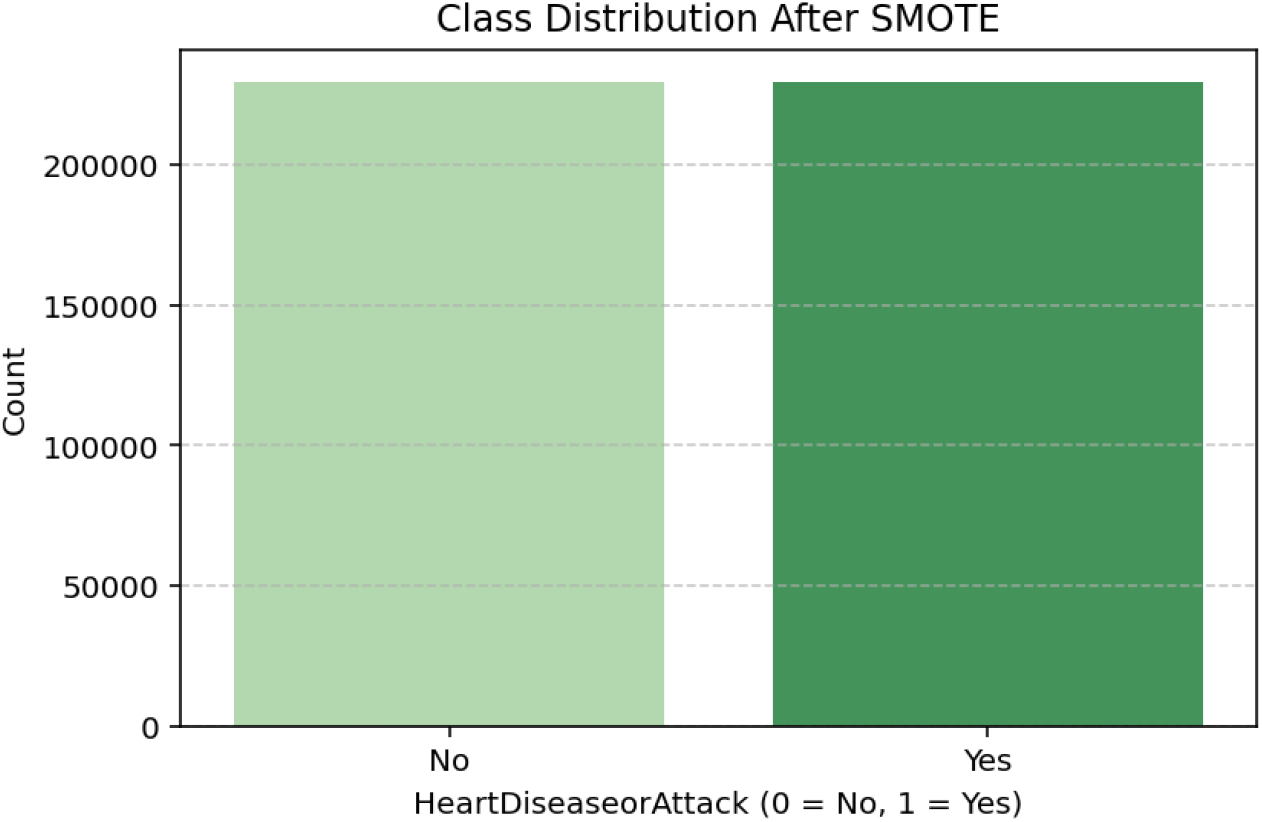
Class Distribution After SMOTE.

Without balancing, models tend to favour the majority class, leading to biased predictions and poor sensitivity in detecting actual positive cases—particularly problematic in a health-related context where correctly identifying high-risk individuals is critical. These visualizations, therefore, reinforce the necessity of techniques like SMOTE in ensuring fair and effective model performance.

### Clustering Analysis

#### Demographic Clusters

The demographic clustering produced eight clusters with a silhouette score of 0.273, indicating moderate cohesion among groups. A chi-square test showed a strong association with CHD (*p <* .001). As shown in Table 3, logistic regression identified Clusters 1 through 3 as high-risk groups, each with odds ratios exceeding 3.75 (*p <* .001). Cluster 6 also presented elevated odds of CHD (OR = 2.03, *p <* .001), while Cluster 5 displayed a modest but statistically significant increase in risk (OR = 1.24, *p* = .027). These patterns suggest that demographic subpopulations characterized by socioeconomic or age-related vulnerabilities are disproportionately affected by CHD. The cluster-wise distribution of CHD cases is illustrated in Figure 4, highlighting the relative burden across subgroups.

**Table 3.** Logistic regression for demographic clusters in predicting CHD.

| Cluster | Coef | Std Err | p-value | OR | Lower CI | Upper CI |
| --- | --- | --- | --- | --- | --- | --- |
| Intercept | -0.836 | 0.070 | ¡ 0.001 | 0.43 | 0.38 | 0.50 |
| Cluster 1 | 1.332 | 0.088 | ¡ 0.001 | 3.79 | 3.19 | 4.50 |
| Cluster 2 | 1.341 | 0.088 | ¡ 0.001 | 3.82 | 3.21 | 4.54 |
| Cluster 3 | 1.346 | 0.088 | ¡ 0.001 | 3.84 | 3.23 | 4.57 |
| Cluster 4 | 0.628 | 0.089 | ¡ 0.001 | 1.87 | 1.57 | 2.23 |
| Cluster 5 | 0.217 | 0.098 | 0.027 | 1.24 | 1.02 | 1.51 |
| Cluster 6 | 0.706 | 0.094 | ¡ 0.001 | 2.03 | 1.69 | 2.43 |
| Cluster 7 | 0.344 | 0.096 | ¡ 0.001 | 1.41 | 1.17 | 1.70 |

**Fig 4.**
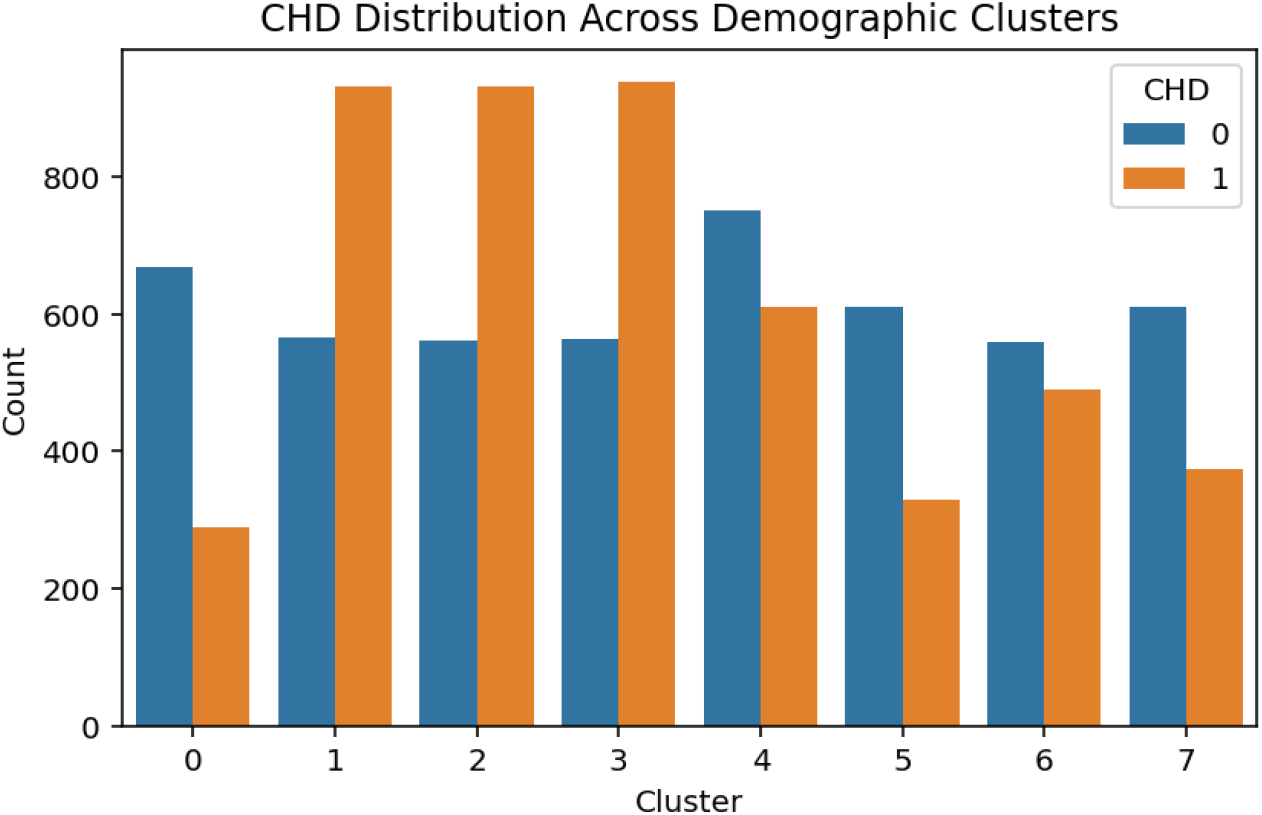
CHD Distribution Across Demographic Clusters

#### Clinical Clusters

Clustering based on clinical characteristics produced nine distinct clusters and yielded a high silhouette score of 0.666, indicating strong structural separation between subgroups. A chi-square test confirmed a statistically significant association between cluster membership and CHD status (*p <* .001). As shown in Table 4, Cluster 5 exhibited an exceptionally high risk of CHD with an odds ratio (OR) of 12.37, followed by Cluster 8 (OR = 4.73), Cluster 7 (OR = 2.42), and Cluster 6 (OR = 2.17). In contrast, Clusters 1 and 4 showed protective associations (ORs *<* 1.0), suggesting these subgroups may contain individuals with fewer or better-managed clinical risk factors. The visual distribution of CHD cases across clusters in Figure 5 further supports these findings, with higher CHD counts clearly concentrated in clusters associated with elevated odds.

**Table 4.** Logistic regression for clinical clusters in predicting CHD.

| Cluster | Coef | Std Err | p-value | OR | Lower CI | Upper CI |
| --- | --- | --- | --- | --- | --- | --- |
| Intercept | 0.009 | 0.054 | 0.872 | 1.01 | 0.91 | 1.12 |
| Cluster 1 | -1.096 | 0.069 | ¡ 0.001 | 0.33 | 0.29 | 0.38 |
| Cluster 2 | 0.343 | 0.091 | ¡ 0.001 | 1.41 | 1.18 | 1.68 |
| Cluster 3 | 0.502 | 0.172 | 0.003 | 1.65 | 1.18 | 2.31 |
| Cluster 4 | -0.649 | 0.078 | ¡ 0.001 | 0.52 | 0.45 | 0.61 |
| Cluster 5 | 2.515 | 0.177 | ¡ 0.001 | 12.37 | 8.74 | 17.49 |
| Cluster 6 | 0.777 | 0.098 | ¡ 0.001 | 2.17 | 1.79 | 2.63 |
| Cluster 7 | 0.886 | 0.085 | ¡ 0.001 | 2.42 | 2.05 | 2.87 |
| Cluster 8 | 1.554 | 0.106 | ¡ 0.001 | 4.73 | 3.84 | 5.82 |

**Fig 5.**
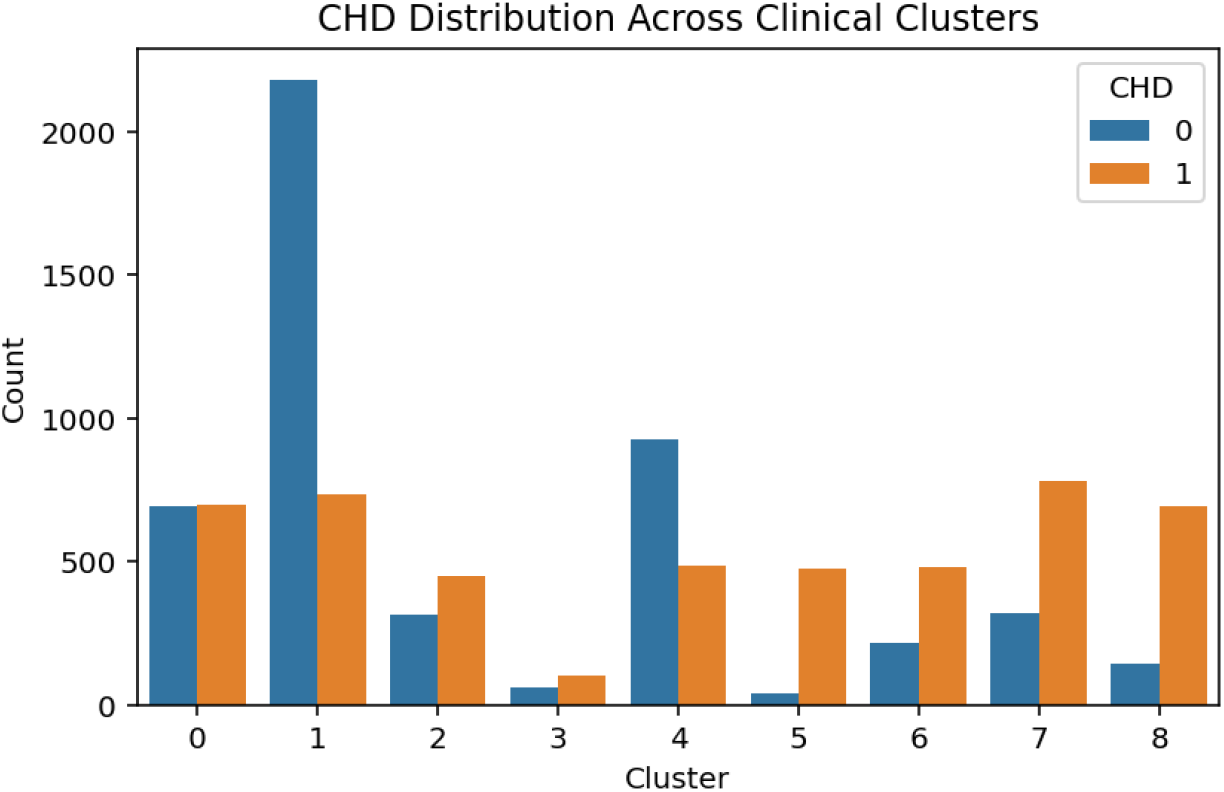
CHD Distribution Across Clinical Clusters

#### Behavioral Clusters

The behavioral clustering resulted in nine distinct groups with a high silhouette score of 0.700, indicating strong internal cohesion and separation between clusters. The chi-square test confirmed a significant association with CHD (*p <* .001). As shown in Table 5, logistic regression revealed that Cluster 2 was associated with significantly elevated odds of CHD (OR = 2.59, *p <* .001). In contrast, Clusters 3, 4, 5, and 8 demonstrated strong protective effects, with odds ratios ranging from 0.07 to 0.17, likely representing behaviorally healthier groups (e.g., non-smokers, physically active individuals). Cluster 1 did not show a statistically significant difference (*p* = .215), suggesting a more neutral behavior-risk profile. These findings are visually supported by Figure 6, which displays the distribution of CHD cases across behavioral clusters.

**Table 5.** Logistic regression for behavioral clusters in predicting CHD.

| Cluster | Coef | Std Err | p-value | OR | Lower CI | Upper CI |
| --- | --- | --- | --- | --- | --- | --- |
| Intercept | 0.744 | 0.066 | ¡ 0.001 | 2.11 | 1.85 | 2.39 |
| Cluster 1 | 0.116 | 0.093 | 0.215 | 1.12 | 0.94 | 1.35 |
| Cluster 2 | 0.950 | 0.090 | ¡ 0.001 | 2.59 | 2.17 | 3.09 |
| Cluster 3 | -2.236 | 0.124 | ¡ 0.001 | 0.11 | 0.08 | 0.14 |
| Cluster 4 | -2.524 | 0.121 | ¡ 0.001 | 0.08 | 0.06 | 0.10 |
| Cluster 5 | -1.799 | 0.090 | ¡ 0.001 | 0.17 | 0.14 | 0.20 |
| Cluster 6 | -0.239 | 0.093 | 0.011 | 0.79 | 0.66 | 0.95 |
| Cluster 7 | -0.784 | 0.096 | ¡ 0.001 | 0.46 | 0.38 | 0.55 |
| Cluster 8 | -2.684 | 0.112 | ¡ 0.001 | 0.07 | 0.05 | 0.09 |

**Fig 6.**
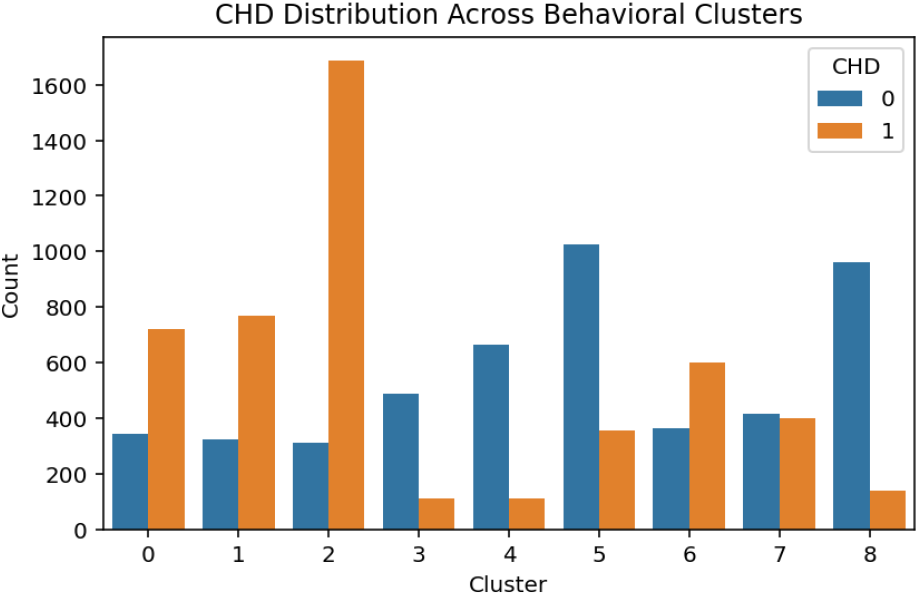
CHD Distribution Across Behavioral Clusters

### Healthcare and Well–being Clusters

Three clusters were identified in the healthcare and well-being domain, with a silhouette score of 0.347. Although the cluster cohesion was lower than in other domains, the association with CHD remained statistically significant (*p <* .001). As shown in Table 6, Cluster 2 exhibited elevated odds of CHD (OR = 1.62, *p <* .001), potentially reflecting individuals facing barriers such as poor access to care, high out-of-pocket costs, or lower self-rated health. In contrast, Cluster 1 showed a strong protective association (OR = 0.28, *p <* .001), likely representing individuals with stable healthcare access and more favorable well-being indicators. These trends are visually corroborated in Figure 7, which displays the distribution of CHD cases across the healthcare clusters.

**Table 6.**
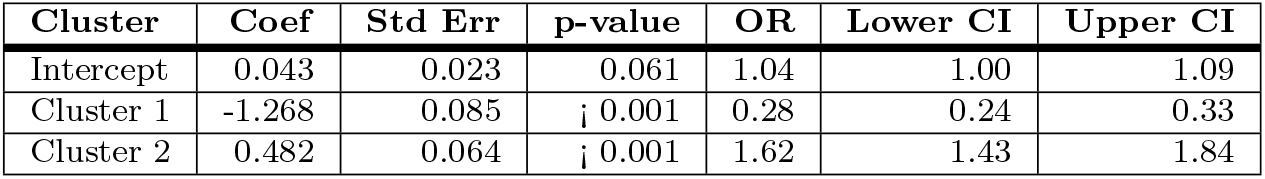
Logistic regression for healthcare clusters in predicting CHD.

| Cluster | Coef | Std Err | p-value | OR | Lower CI | Upper CI |
| --- | --- | --- | --- | --- | --- | --- |
| Intercept | 0.043 | 0.023 | 0.061 | 1.04 | 1.00 | 1.09 |
| Cluster 1 | -1.268 | 0.085 | ¡ 0.001 | 0.28 | 0.24 | 0.33 |
| Cluster 2 | 0.482 | 0.064 | ¡ 0.001 | 1.62 | 1.43 | 1.84 |

**Fig 7.**
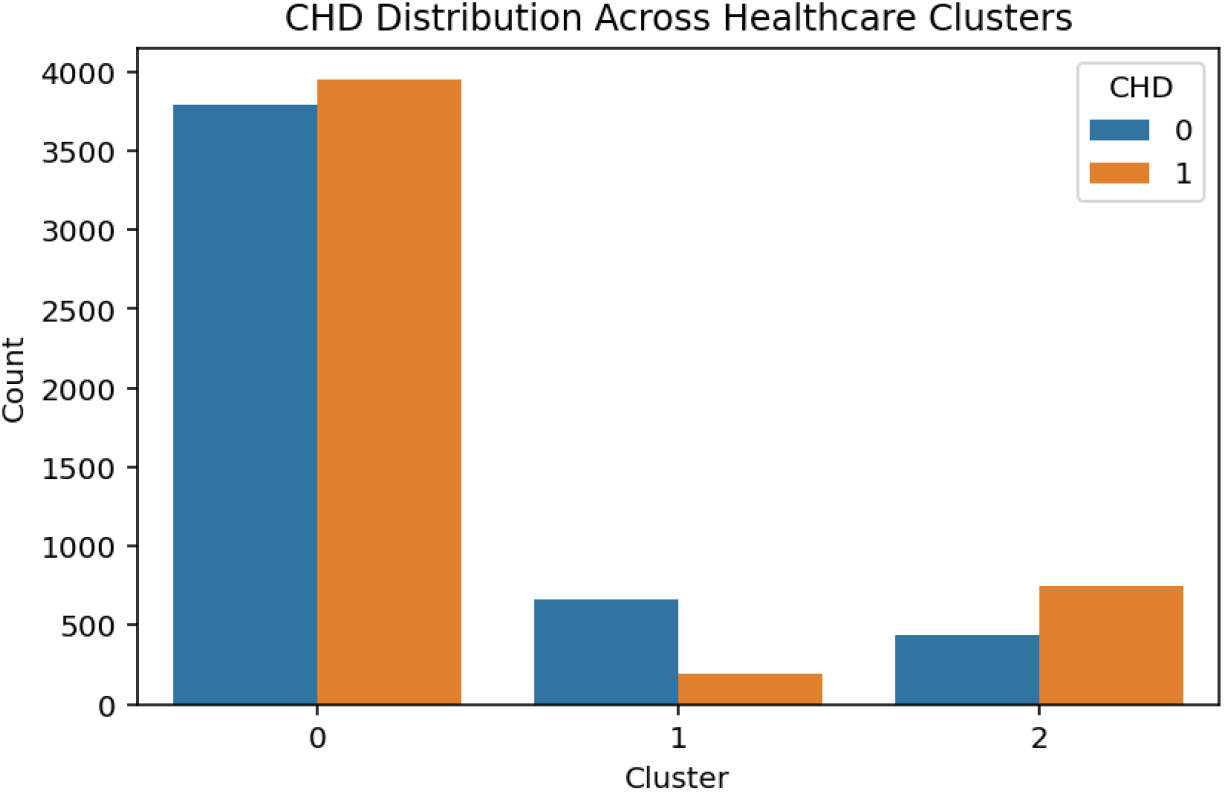
CHD Distribution Across Healthcare Clusters

### Full Feature Clusters

Clustering across the full feature set resulted in five integrated clusters, though the silhouette score was relatively low (0.077), indicating weak separation among clusters. Nevertheless, the association with CHD remained highly significant (*p <* .001). As presented in Table 7, Cluster 2 exhibited exceptionally high odds of CHD (OR = 24.36), followed by Clusters 1 and 4 (ORs = 7.32 and 7.78, respectively). These clusters likely capture individuals with combined demographic, clinical, and behavioral vulnerabilities. In contrast, Cluster 3 had significantly lower odds of CHD (OR = 0.70, *p* = .004), suggesting the presence of a potentially resilient subgroup. Despite the reduced silhouette cohesion, this integrated domain provided valuable insight into the overlapping risk factors influencing CHD. These findings are visually represented in Figure 8, which shows the distribution of CHD cases across the full feature clusters.

**Table 7.** Logistic regression results for full feature clusters predicting coronary heart disease (CHD).

| Cluster | Coef | Std Err | p-value | OR | Lower CI | Upper CI |
| --- | --- | --- | --- | --- | --- | --- |
| Intercept | -1.541 | 0.109 | ¡ 0.001 | 0.21 | 0.17 | 0.26 |
| Cluster 1 | 1.991 | 0.113 | ¡ 0.001 | 7.32 | 5.86 | 9.14 |
| Cluster 2 | 3.193 | 0.123 | ¡ 0.001 | 24.36 | 19.15 | 30.99 |
| Cluster 3 | -0.353 | 0.122 | 0.004 | 0.70 | 0.55 | 0.89 |
| Cluster 4 | 2.052 | 0.196 | ¡ 0.001 | 7.78 | 5.30 | 11.43 |

**Table 8.**
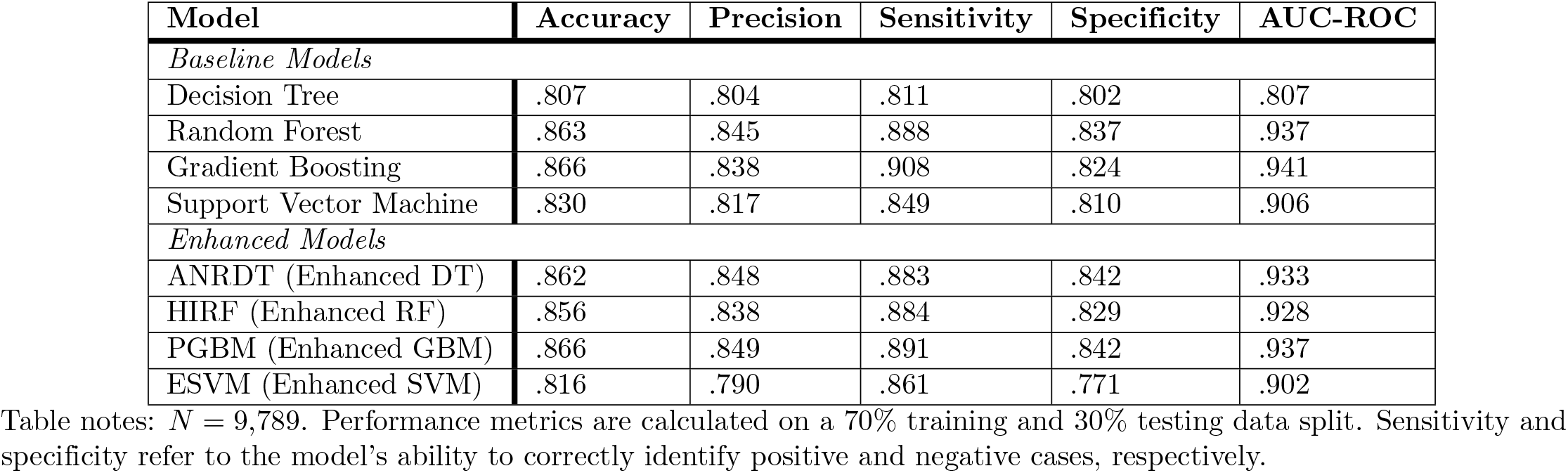
Comparison of baseline and enhanced machine learning models on classification metrics.

| Model | Accuracy | Precision | Sensitivity | Specificity | AUC-ROC |
| --- | --- | --- | --- | --- | --- |
| *Baseline Models* |  |  |  |  |  |
| Decision Tree | .807 | .804 | .811 | .802 | .807 |
| Random Forest | .863 | .845 | .888 | .837 | .937 |
| Gradient Boosting | .866 | .838 | .908 | .824 | .941 |
| Support Vector Machine | .830 | .817 | .849 | .810 | .906 |
| *Enhanced Models* |  |  |  |  |  |
| ANRDT (Enhanced DT) | .862 | .848 | .883 | .842 | .933 |
| HIRF (Enhanced RF) | .856 | .838 | .884 | .829 | .928 |
| PGBM (Enhanced GBM) | .866 | .849 | .891 | .842 | .937 |
| ESVM (Enhanced SVM) | .816 | .790 | .861 | .771 | .902 |

Table notes: $N = 9,789$ . Performance metrics are calculated on a 70% training and 30% testing data split. Sensitivity and specificity refer to the model's ability to correctly identify positive and negative cases, respectively.

**Fig 8.**
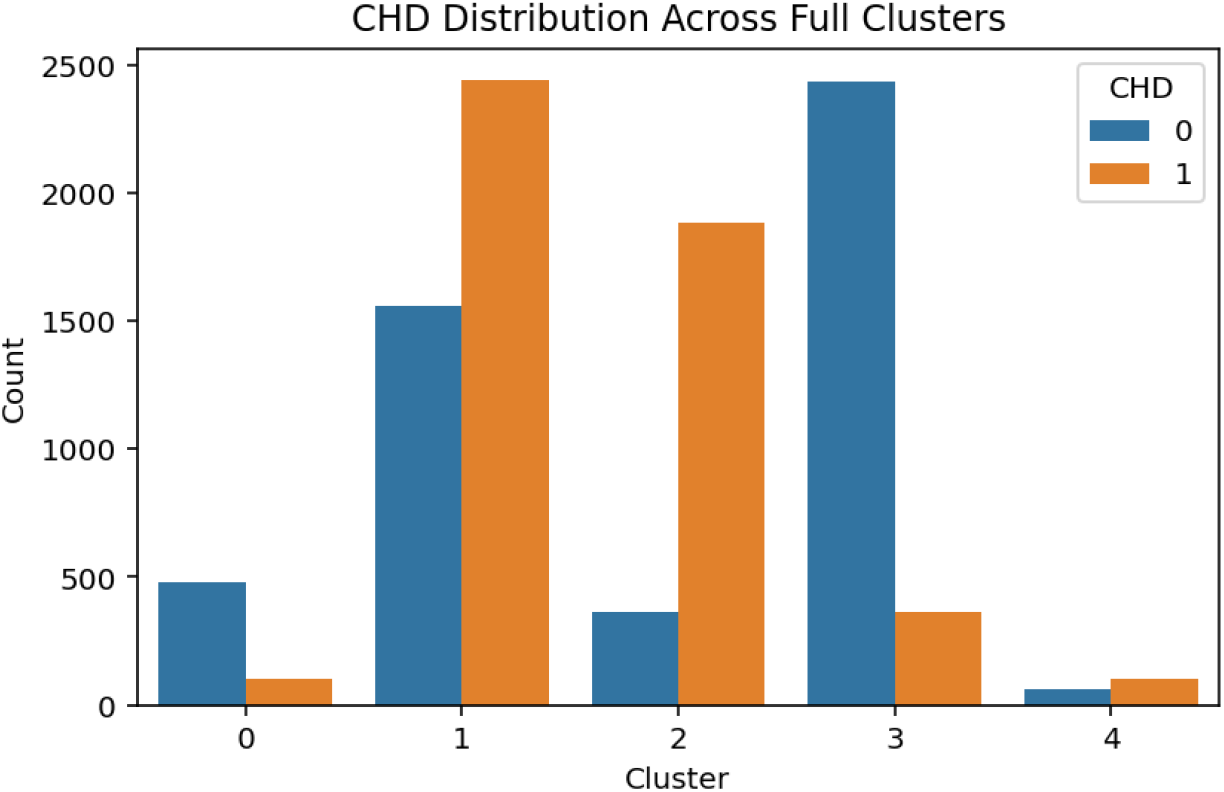
CHD Distribution Across Full Feature Clusters

### Confused Matrix of Baseline Ensemble Models

#### Bayesian Model Averaging (BMA)

Figure 9 displays the confusion matrix for the baseline Bayesian Model Averaging (BMA) model. This method combines probabilistic predictions from various base classifiers using Bayesian principles to produce an averaged output. The model correctly classified 839 non–CHD cases (true negatives) and 876 CHD-positive cases (true positives). However, it also misclassified 161 nonC–HD instances as CHD (false positives) and failed to detect 124 true CHD cases (false negatives). The model’s sensitivity is calculated as:

**Fig 9.**
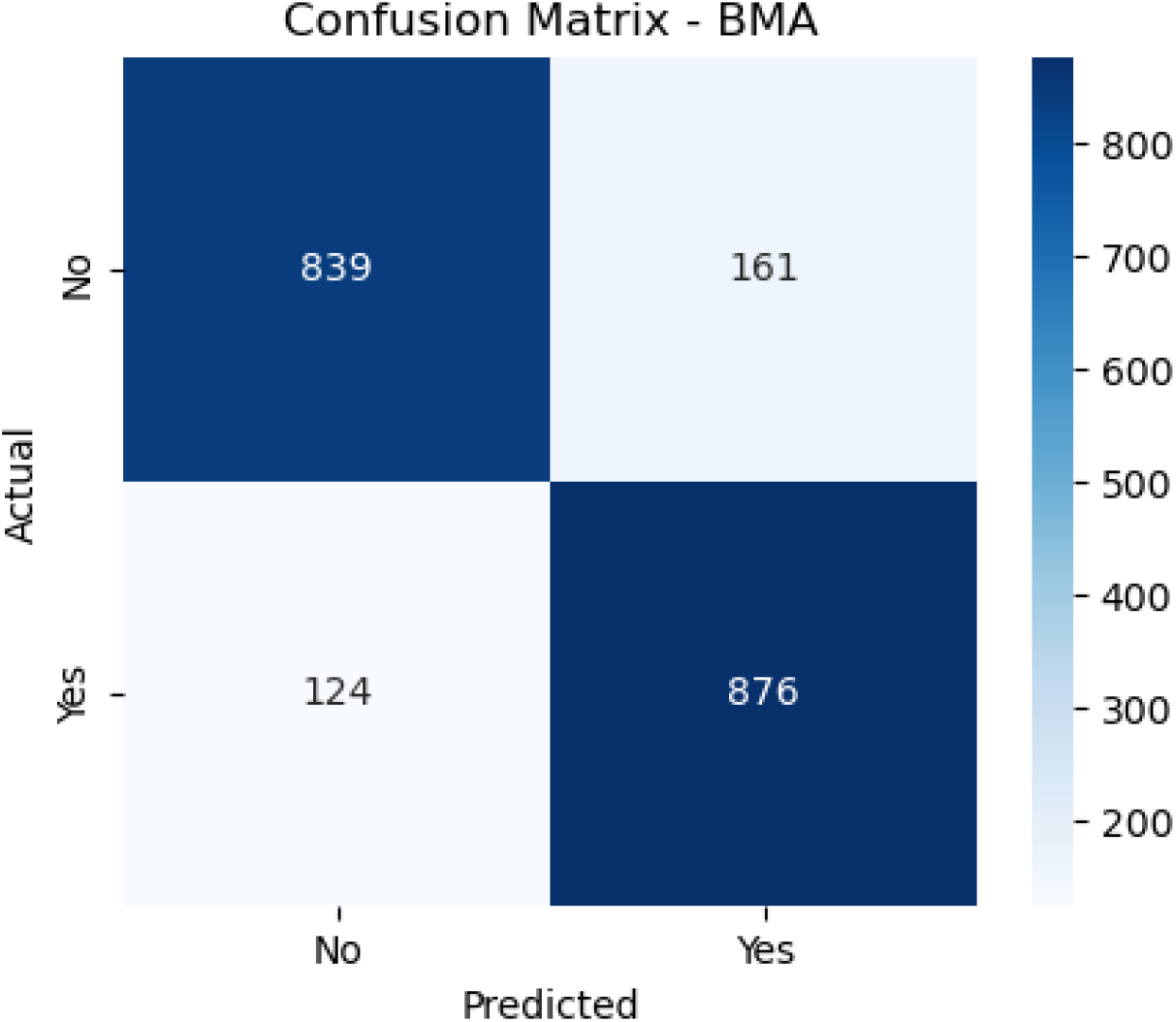
Confusion Matrix for Bayesian Model Averaging (BMA)

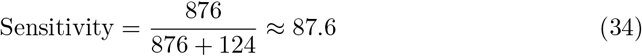

which shows its ability to correctly identify patients with CHD. The specificity, representing its performance on the negative class, is:

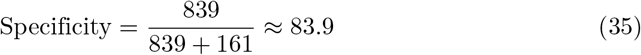

These values suggest that while BMA performs reasonably well in identifying CHD-positive individuals, it has a moderate rate of false alarms. This trade-off is expected due to the averaging nature of the method, which might smooth over sharp decision boundaries.

#### Majority Voting

Figure 10 illustrates the confusion matrix for the baseline Majority Voting ensemble, a straightforward yet robust method where each base learner casts a vote and the majority prediction is selected. The model achieved 854 true negatives and 873 true positives. It also misclassified 146 non-CHD patients as positive (false positives) and missed 127 CHD cases (false negatives). The sensitivity is given by:

**Fig 10.**
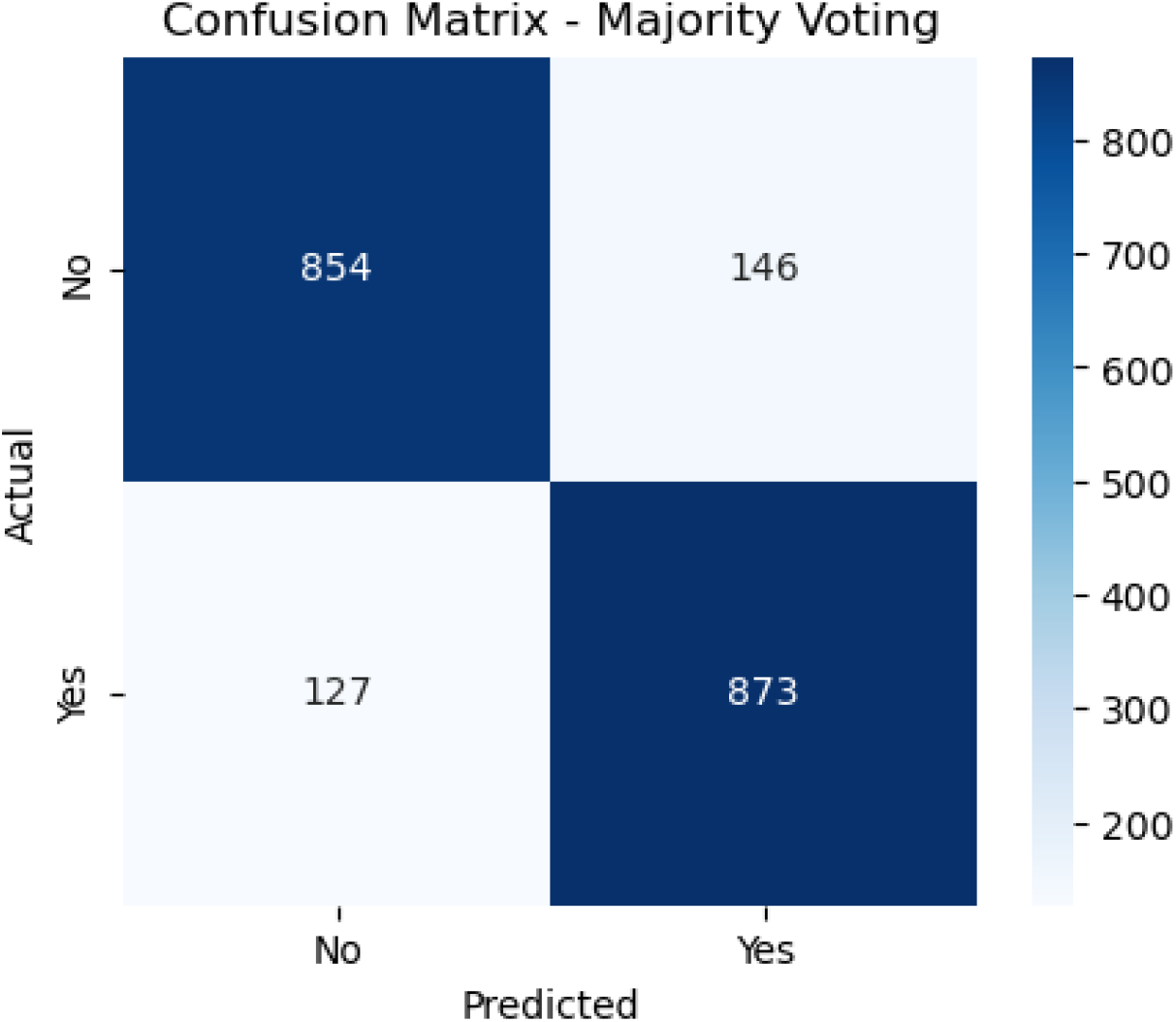
Confusion Matrix for Majority Voting

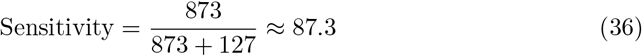

and the specificity is:

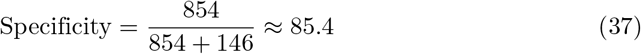

These results suggest that Majority Voting strikes a decent balance between identifying CHD-positive individuals and minimizing false positives. However, due to the equal weighting of all classifiers, it may overlook the strengths of stronger learners in certain cases.

#### Boosting

Figure 11 presents the confusion matrix for the baseline Boosting model, which trains weak learners sequentially while correcting previous errors. The model correctly identified 908 CHD-positive patients (true positives) and 824 non-CHD individuals (true negatives). However, it misclassified 92 CHD patients (false negatives) and 176 non–CHD patients (false positives). Sensitivity is computed as:

**Fig 11.**
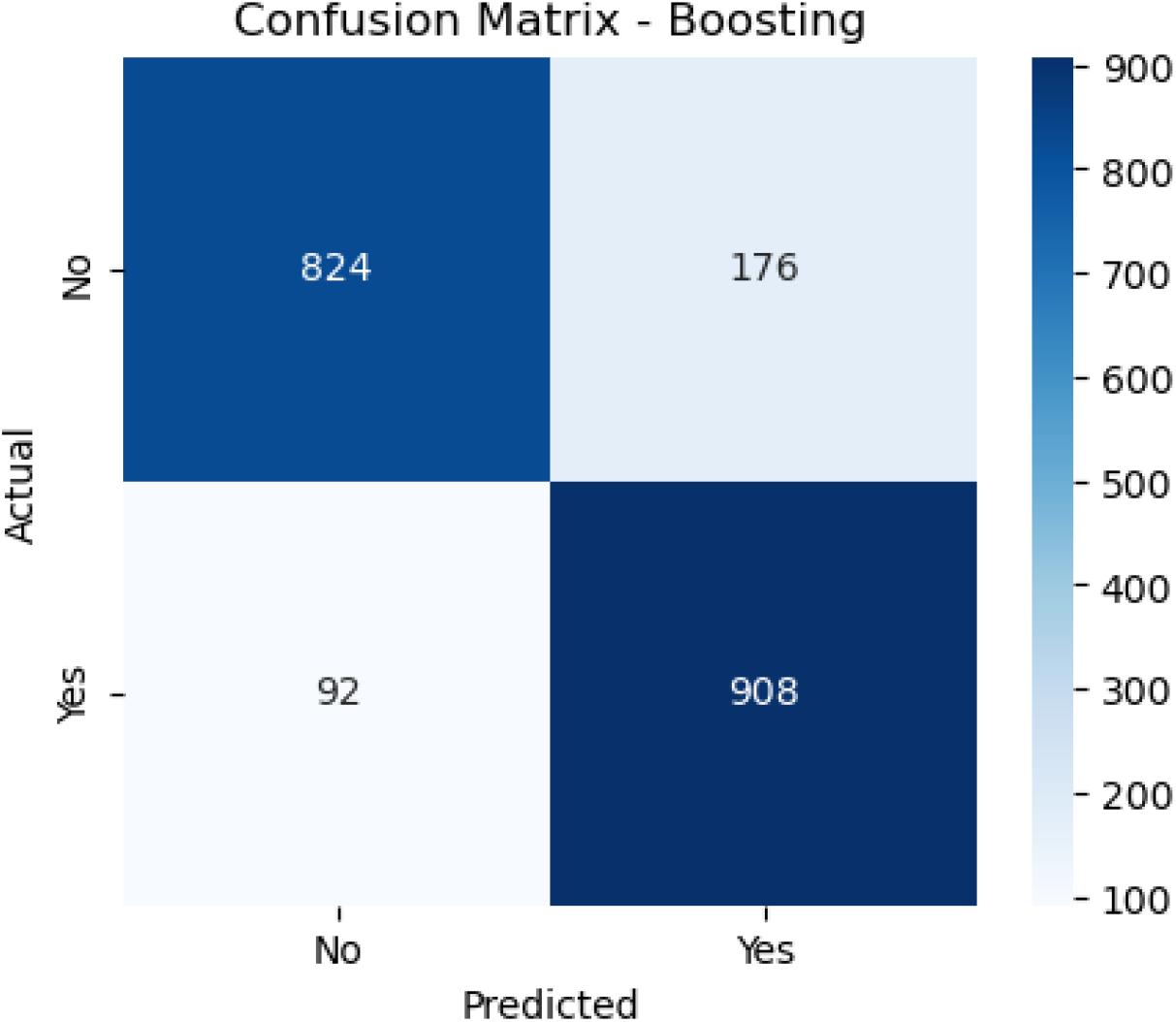
Confusion Matrix for Boosting (Baseline)

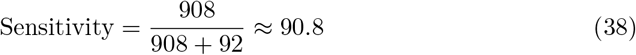

indicating excellent performance in detecting CHD. Specificity is:

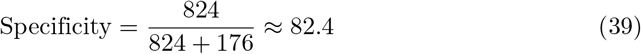

This shows that while Boosting is aggressive in minimizing false negatives (which is critical in medical diagnosis), it incurs a slightly higher false positive rate. The high sensitivity makes Boosting particularly effective in clinical scenarios requiring thorough screening.

#### Bagging

Figure 12, illustrates the confusion matrix for the Bagging model—based on bootstrap aggregation—achieved 830 true negatives and 882 true positives. It recorded 170 false positives and 118 false negatives. The sensitivity of the Bagging model is:

**Fig 12.**
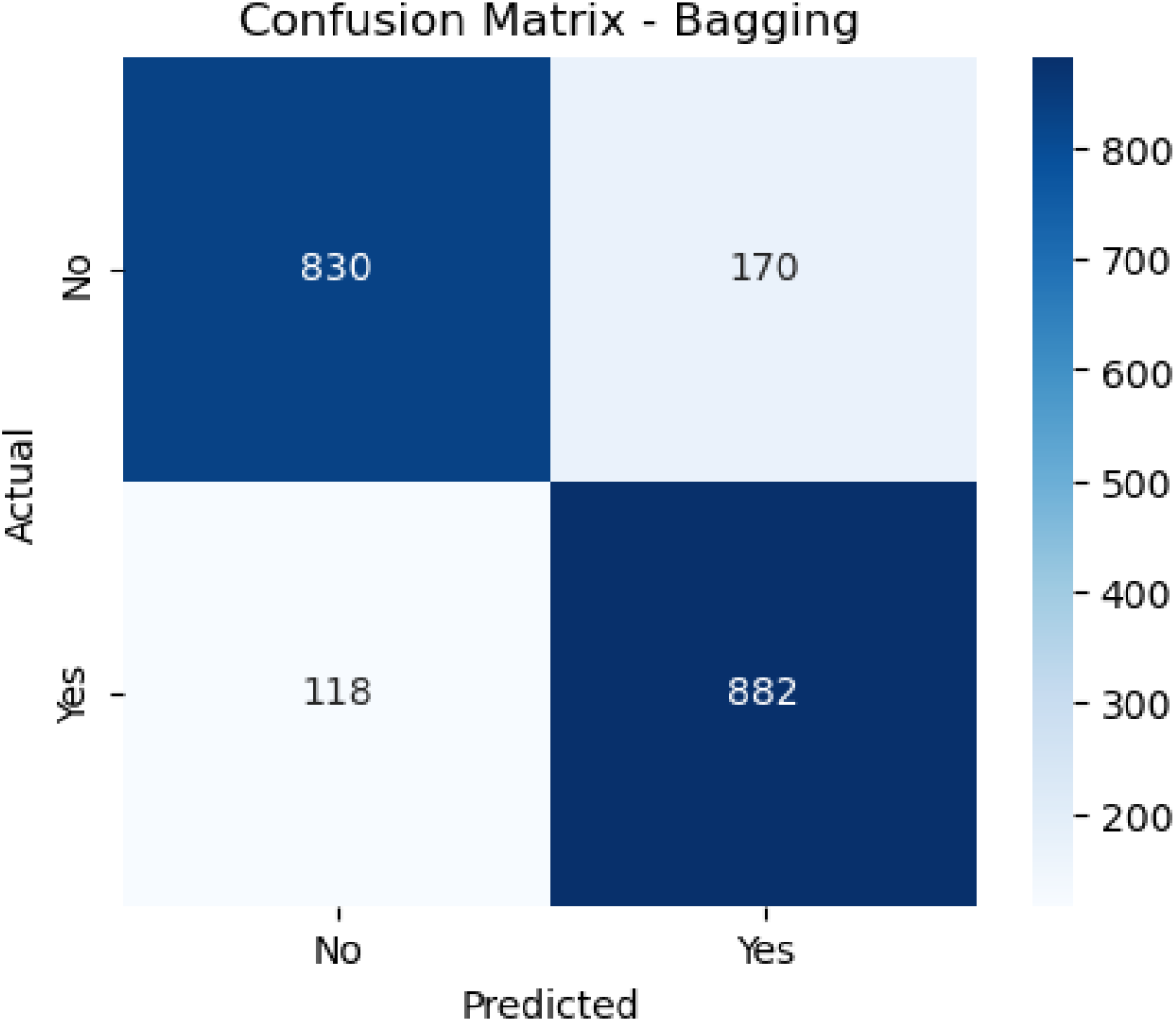
Confusion Matrix for Bagging (Baseline)

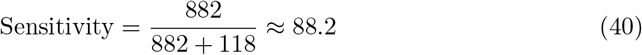

and specificity is:

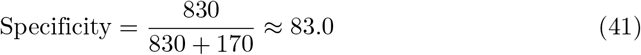

Bagging shows a well–balanced trade–off between both classes. Although not as aggressive as Boosting, it is generally more robust to variance, offering reliable performance across different data samples, which is advantageous in real–world medical datasets.

#### Stacking

Figure 13 shows the performance of the baseline Stacking ensemble, which uses a meta–learner to combine predictions from multiple base models. The stacking model correctly predicted 846 non–CHD cases (true negatives) and 898 CHD-positive cases (true positives). It also reported 154 false positives and 102 false negatives. The sensitivity is:

**Fig 13.**
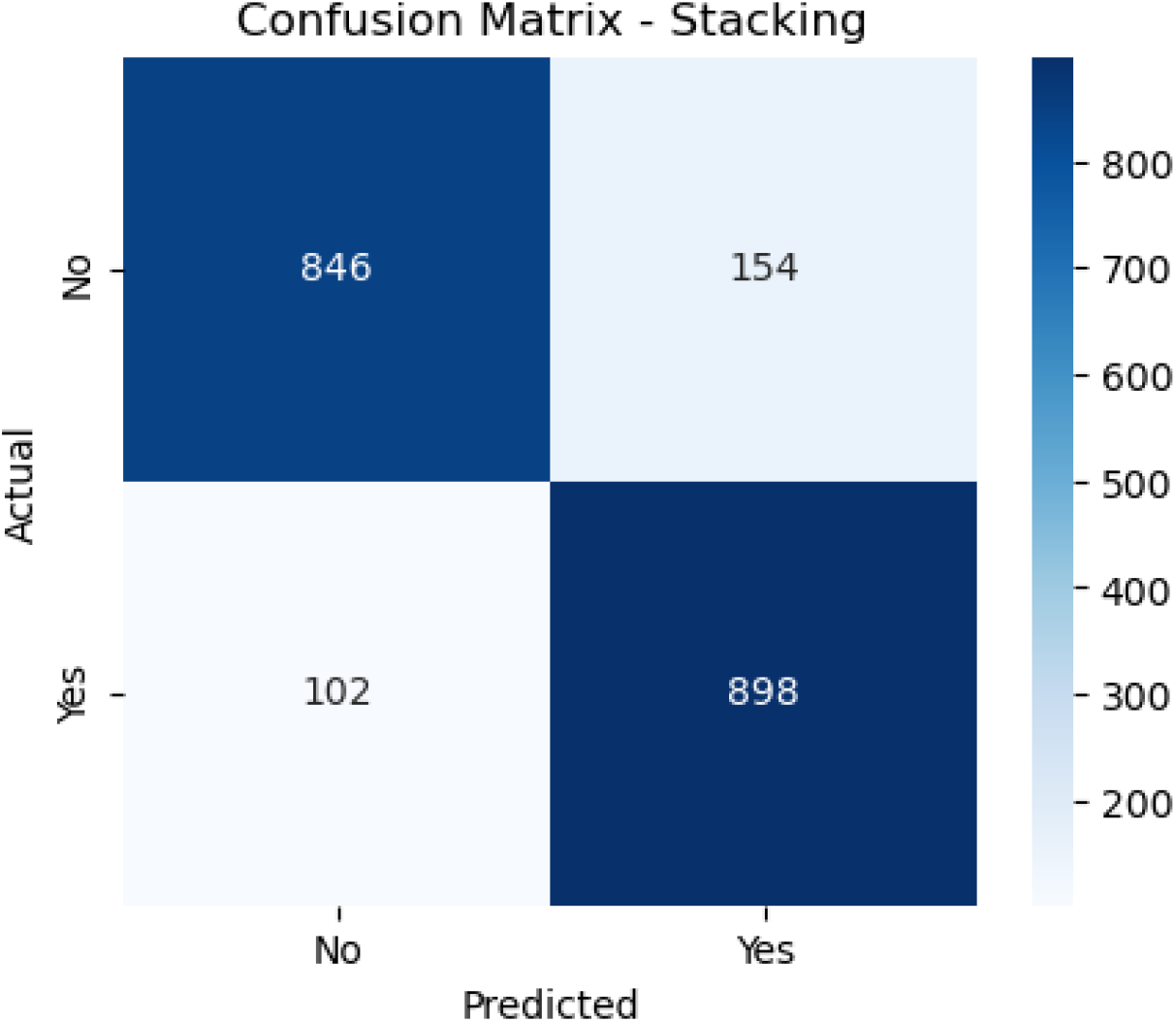
Confusion Matrix for Stacking (Baseline)

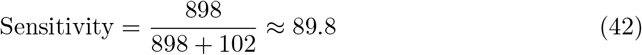

and the specificity is:

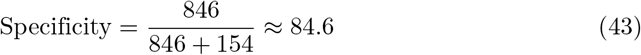

These metrics indicate strong and stable performance, driven by the meta-learner’s capacity to synthesize multiple decision boundaries. Stacking proves effective at improving the strengths of individual models, offering high sensitivity for clinical needs and strong specificity for reducing overdiagnosis.

### Confused Matrix of Enhanced/Improved Ensemble Models

#### Bayesian Model Averaging (BMA)

The confusion matrix on Figure 14 illustrates the performance of the Bayesian Model Averaging (BMA) ensemble method. Out of all actual negative cases (i.e., patients who did not have CHD), the model correctly predicted 821 as negative (true negatives) and misclassified 158 as positive (false positives). Conversely, among the actual positive cases (patients with CHD), it correctly identified 867 as positive (true positives), while misclassifying 112 as negative (false negatives).

**Fig 14.**
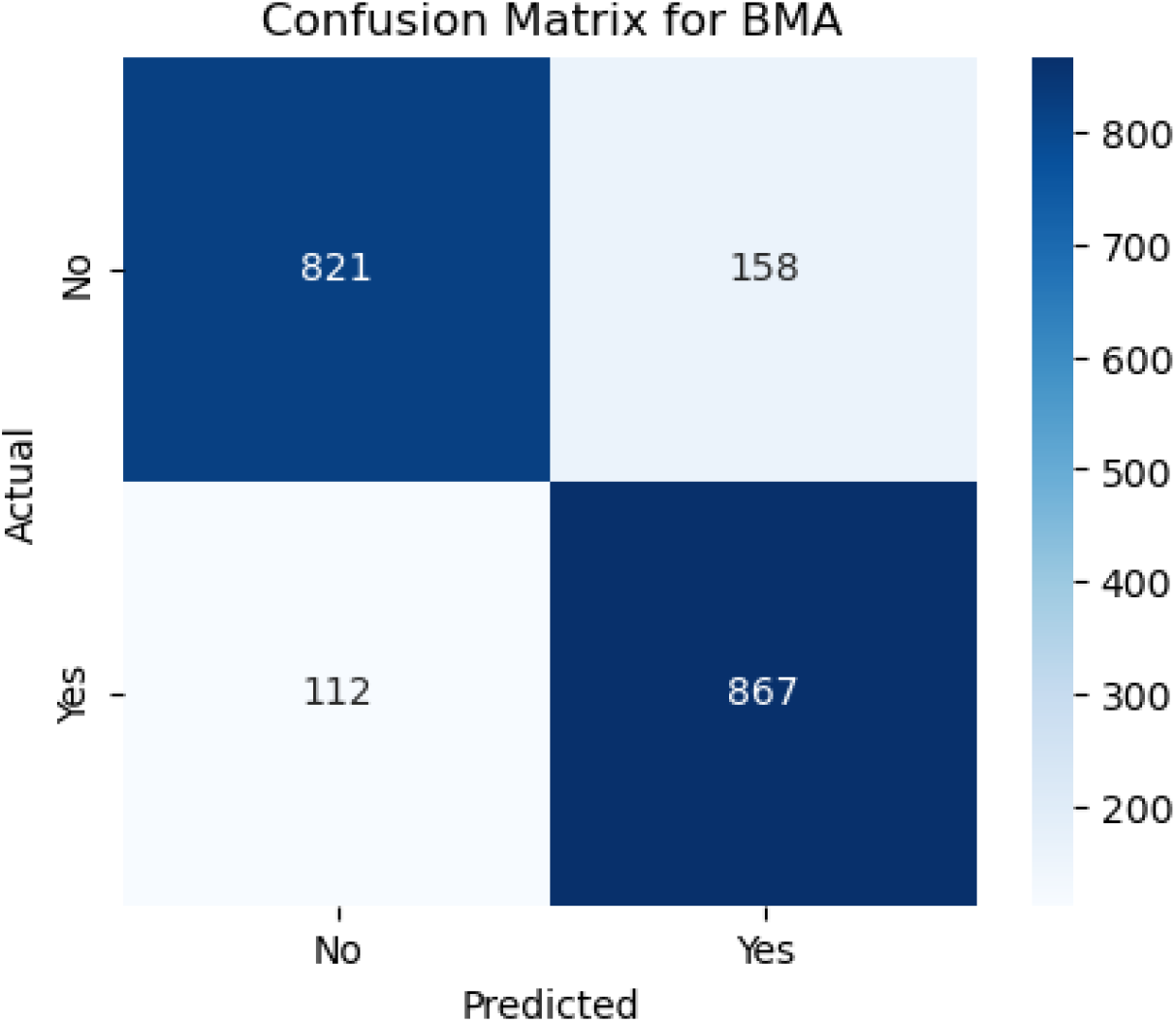
Confusion Matrix for Bayessian Model Averaging Ensemble

These results translate into strong diagnostic performance, with the BMA model demonstrating high sensitivity (recall) and strong specificity. Sensitivity is calculated as:

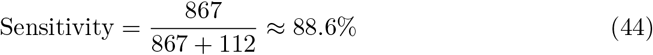

indicating the model’s excellent ability to identify patients who truly have CHD. Specificity is:

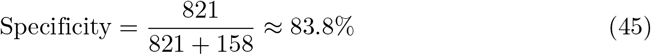

reflecting its capacity to correctly dismiss non–CHD cases. However, the presence of 158 false positives might result in unnecessary further testing or anxiety for patients. On the other hand, the false negative count of 112 is comparatively low, which is crucial in clinical settings where missing true CHD cases can have severe consequences. Overall, the BMA ensemble model offers a reliable balance between minimizing both false negatives and false positives.

#### Majority Voting

Figure 15 displays the confusion matrix for the Majority Voting ensemble method. For actual non–CHD cases, this model correctly predicted 836 as negative (true negatives) and misclassified 143 as CHD (false positives). For actual CHD cases, 852 were accurately identified as positive (true positives), while 127 were misclassified as negative (false negatives).

**Fig 15.**
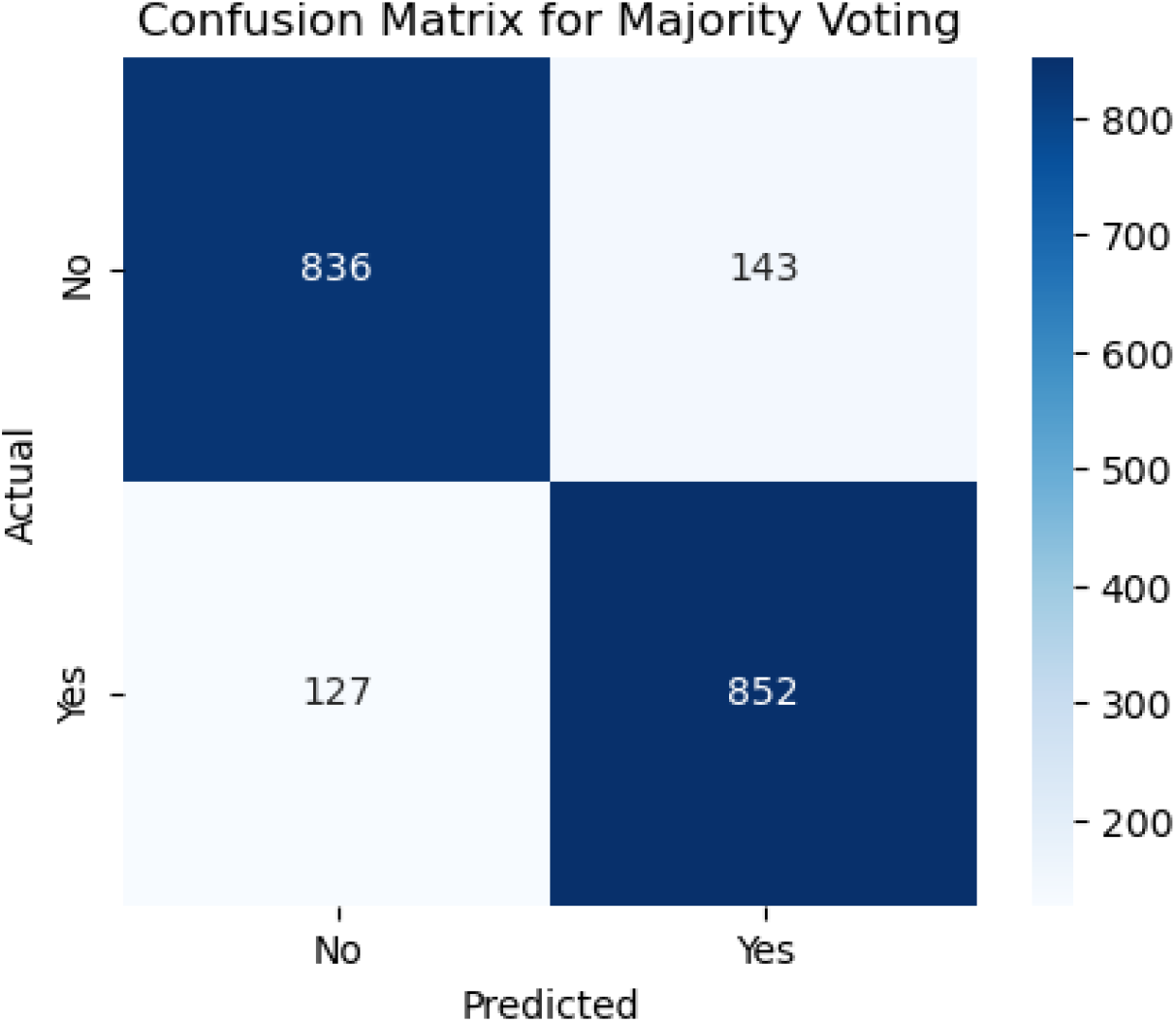
Confusion Matrix for Bayessian Model Averaging Ensemble

Compared to BMA, the Majority Voting model achieved a slightly better specificity: 836

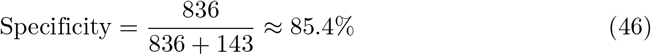

meaning it was marginally more effective at identifying non-CHD cases correctly. However, its sensitivity was lower:

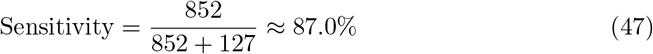

suggesting it missed slightly more true CHD cases than BMA. While this trade–off led to fewer false positives, it slightly increased the risk of failing to detect CHD in patients who actually have the condition. Clinically, this might be less favorable in high-risk populations where identifying every CHD case is a priority, even at the expense of more false positives.

#### Bagging

The confusion matrix on Figure 16 shows the performance of the Bagging (Bootstrap Aggregation) ensemble model. Out of all actual negative cases (non–CHD), 812 were correctly classified as negative (true negatives), while 167 were misclassified as positive (false positives). Among actual positive CHD cases, 865 were correctly identified (true positives), and 114 were misclassified as negative (false negatives).

**Fig 16.**
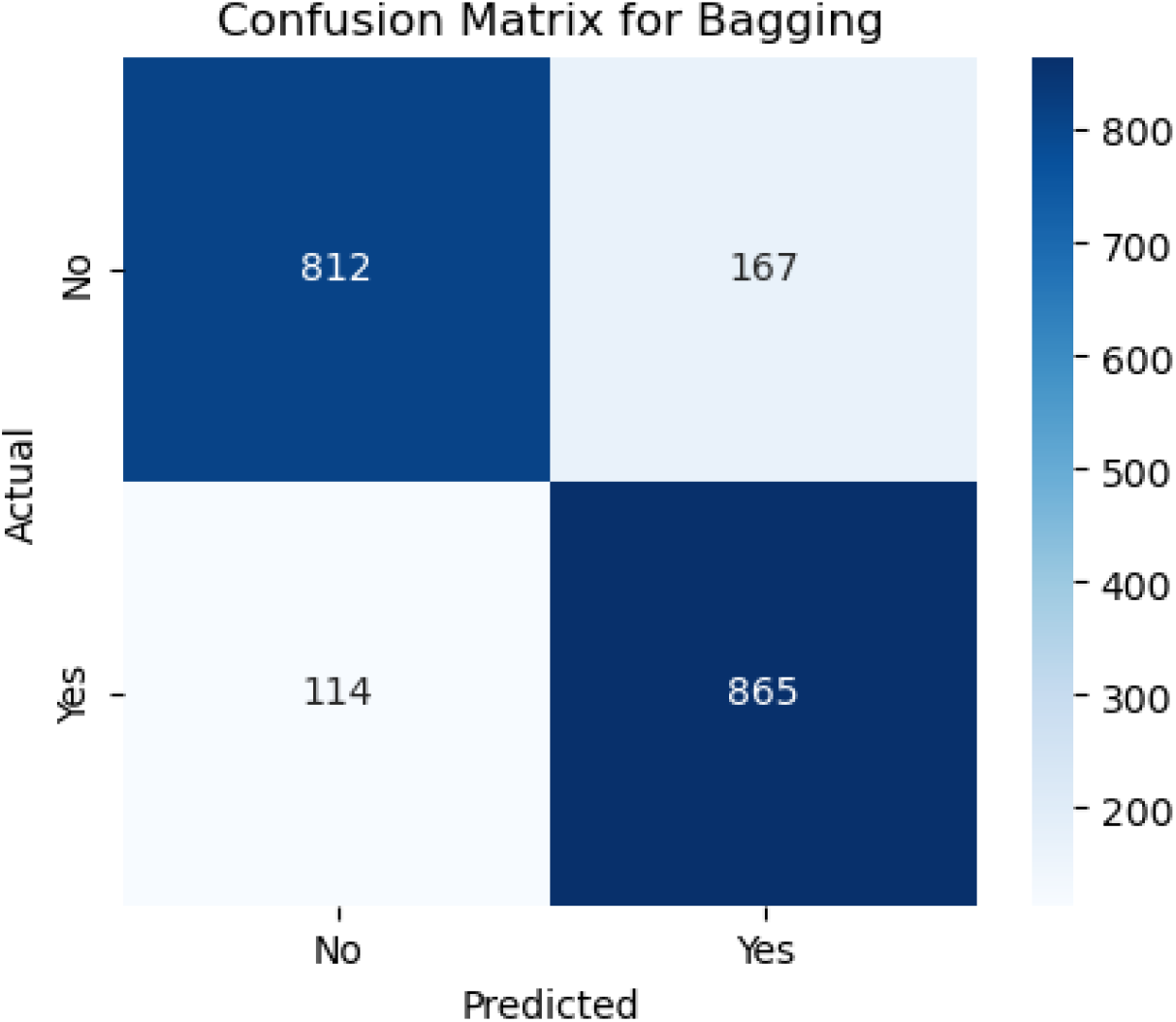
Confusion Matrix for Bagging Ensemble

The sensitivity (recall), calculated as:

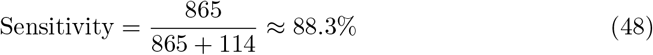

indicates a strong ability to detect CHD cases. The specificity is given by:

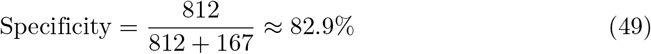

This shows that the model performs well in ruling out non–CHD cases but does produce a fair number of false alarms. Bagging’s effectiveness stems from its strategy of training multiple base learners on different bootstrap samples and averaging their predictions, which reduces variance and improves generalization.

However, the relatively higher false positive count (167) may lead to over-screening or unnecessary clinical follow–up for patients wrongly flagged as at-risk. Despite this, the model maintains a good balance between sensitivity and specificity, making it suitable for applications where slightly higher false positives are acceptable in exchange for fewer missed CHD diagnoses.

#### 0.0.1 Boosting

Figure 17 shows the confusion matrix for the Boosting model, which delivers enhanced performance through sequential learning. The model correctly classified 824 of the non–CHD cases (true negatives) and incorrectly flagged 155 as positive (false positives). It also correctly detected 872 true CHD cases (true positives) and missed 107 positive cases (false negatives).

**Fig 17.**
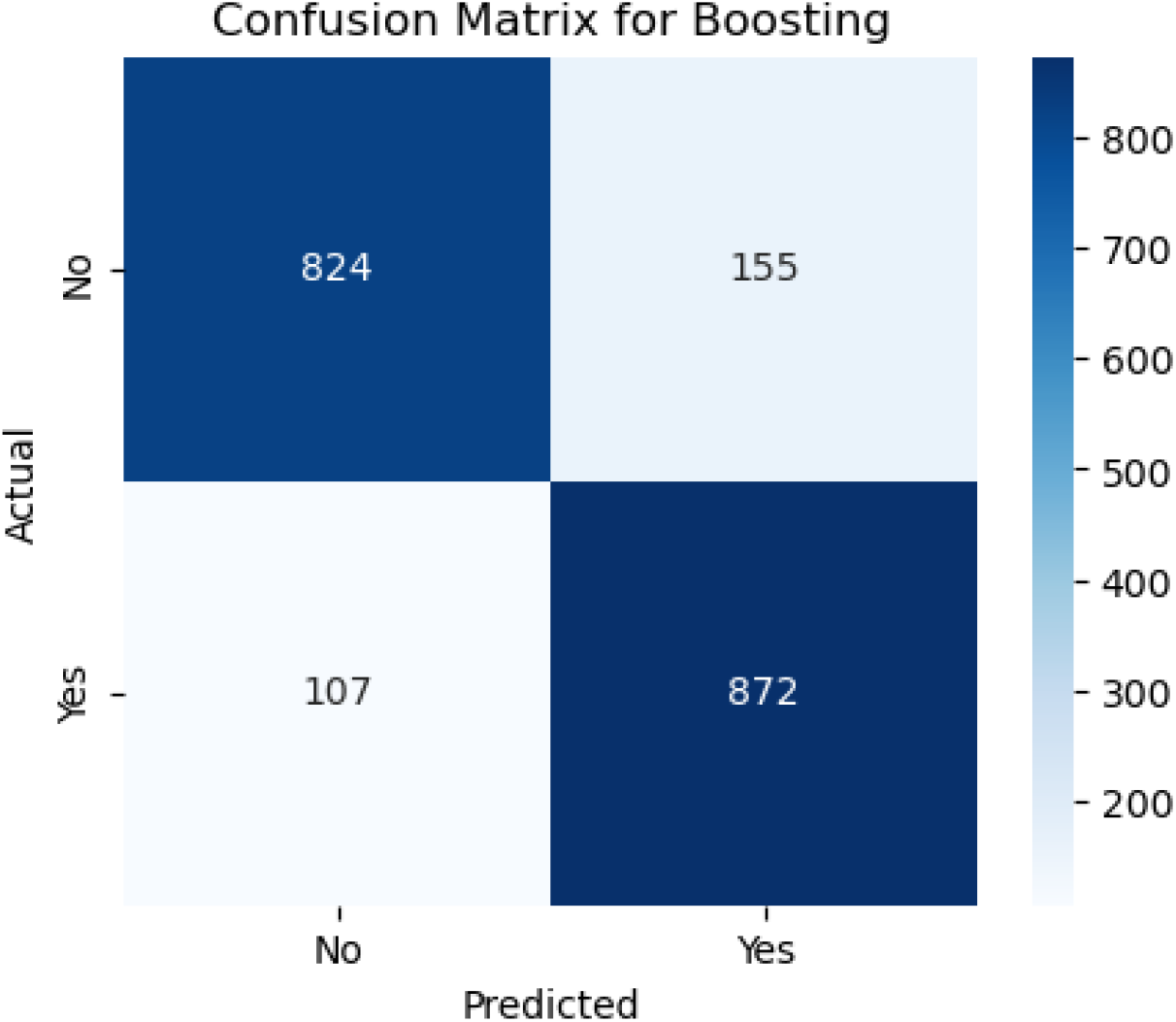
Confusion Matrix for Boosting Ensemble

The sensitivity of the Boosting model is:

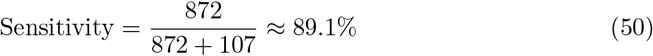

slightly higher than Bagging, suggesting improved detection of CHD-positive patients. The specificity is:

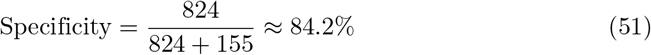

also modestly better than Bagging’s performance. These improved results confirm that Boosting, particularly models like Gradient Boosting or XGBoost, excels in reducing bias by focusing on the mistakes of previous learners during training iterations. The combination of higher sensitivity and specificity makes Boosting highly attractive for clinical screening scenarios where both early detection and minimizing overdiagnosis are critical. Furthermore, the relatively lower false negative count (107) compared to Bagging (114) further reinforces its utility in predictive healthcare applications.

#### Stacking Ensemble Model

The confusion matrix shown in Figure 18 represents the classification performance of the stacking ensemble model, which emerged as the best-performing model in this study. From the matrix, the model correctly predicted 827 of the actual negative cases (true negatives) and misclassified 152 as positive (false positives). In the positive class (patients with CHD), the model correctly identified 872 as true positives and incorrectly labeled 107 cases as negatives (false negatives).

**Fig 18.**
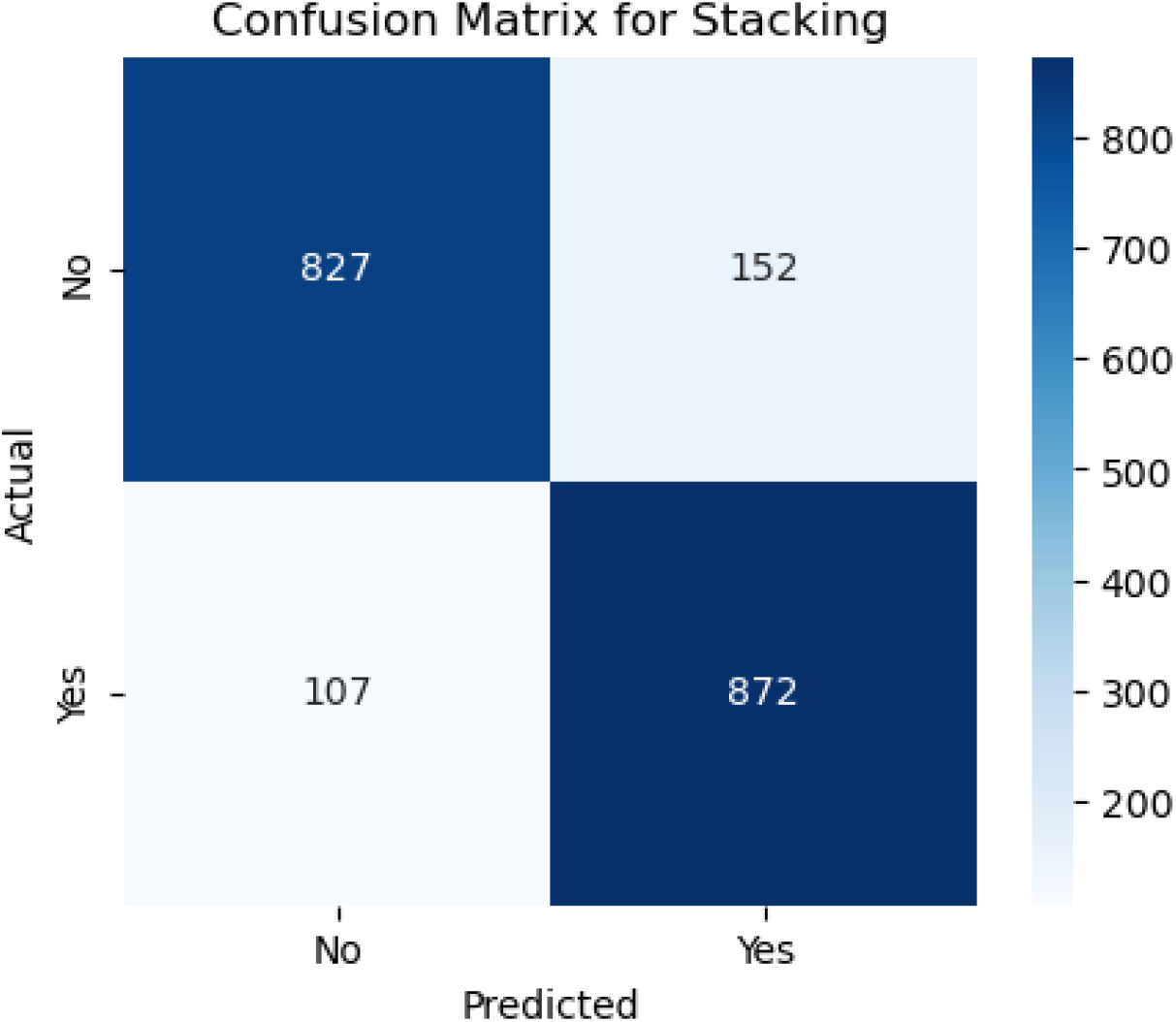
Confusion Matrix for Stacking Ensemble

These results reflect a high sensitivity (recall):

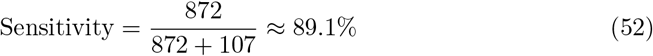

indicating that the stacking model is highly effective at correctly identifying CHD-positive individuals. The specificity, calculated as:

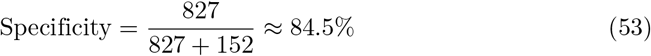

is also quite strong, suggesting good performance in recognizing true negative cases.

These two metrics in tandem confirm that the model maintains a well-balanced classification boundary between the positive and negative classes—crucial in CHD screening, where both under-diagnosis and over-diagnosis can carry significant clinical risks. The relatively low number of false negatives (107) reinforces the model’s suitability for early detection scenarios, while the false positive count (152) remains within an acceptable range for follow-up diagnostic assessment in clinical settings.

The stacking model benefits from its design, which integrates multiple base learners (e.g., logistic regression, random forest, SVM) into a meta-classifier that learns to correct individual model weaknesses. This fusion allows the model to capture both linear and non-linear relationships in the data and leverage complementary decision boundaries from diverse algorithms. The high performance observed in this matrix confirms that stacking not only improves raw accuracy but also optimizes the sensitivity-specificity trade–off better than most standalone models or simpler ensembles like bagging or majority voting.

### Evaluating ML Models: Individual (Baseline) vs Improved Models for Predicting CHD

Individual (baseline) and enhanced ML models were evaluated for predicting CHD–this study utilized a refined and balanced dataset consisting of 9,789 observations obtained after applying SMOTE and stratified random sampling. The data was split using a 70% training and 30% testing ratio, and the models’ predictive performances were assessed using five key classification metrics: Accuracy, Precision, Sensitivity (Recall), Specificity, and AUC-ROC. Both baseline ML models (Decision Tree, Random Forest, Gradient Boosting, and SVM) and improved ML models (ANRDT, HIRF, PGBM, and ESVM) were evaluated to determine how algorithmic enhancements influence predictive accuracy and robustness in the context of CHD.

### Baseline Machine Learning Models

Gradient Boosting (GBM) and Random Forest (RF) emerged as top performers among the baseline models. GBM achieved the highest sensitivity (0.908), indicating a superior ability to identify true positives (patients with CHD), while RF demonstrated a slightly higher AUC-ROC (0.937), denoting excellent discriminative capability. Both models outperformed Decision Tree (DT) and Support Vector Machine (SVM) across nearly all metrics. The Decision Tree, though interpretable, yielded relatively modest performance with an accuracy of 0.807 and a specificity of 0.802, reflecting a limited generalization capacity. While achieving an accuracy of 0.830 and a high AUC-ROC of 0.906, SVM fell short compared to the ensemble approaches in capturing complex patterns in the data.

### Enhanced Machine Learning Models

The enhanced models showed how specific changes proved effective in overcoming the shortcomings of their original versions. Interestingly, the Pruned Gradient Boosting Machine (PGBM) was found to be equivalent to the baseline GBM by accuracy (0.866). It outperformed precision (0.849) and specificity (0.842), showing more balanced performance, fewer false positives, and better generalization. The Hybrid Imbalanced Random Forest (HIRF), based on SMOTE and SHAP values, had a marginally lower overall precision (0.856) but had a better balance of sensitivity (0.884) and specificity (0.829), indicating its effectiveness in class imbalance.

Likewise, the Adaptive Noise-Resistant Decision Tree (ANRDT) has shown significant improvement compared to the baseline decision trees (DT), with improvements seen in all the metrics, namely, accuracy (0.862), sensitivity (0.883), and AUC-ROC (0.933). These improvements bear out the hypothesis that probabilistic splits and regularization lowered overfitting and made them more robust to noisy inputs. Although the Enhanced SVM (ESVM) showed an increase in sensitivity (0.861) and the interpretability of the SHAP integration, it did not perform better than the baseline SVM across all measures. This finding can be attributed to the sensitivity of ESVM to the choice of a kernel and lower specificity (0.771), signalling the difficulty of achieving trade-offs in SVM tuning.

In general, the improved models performed as well as or better than the base ones in all crucial measures, particularly in sensitivity and AUC-ROC, which are essential in the detection of CHD cases. Pruning, reweighting and noise resistance are all ensemble-based improvement methods that offer indicated generalizability, robustness and interpretability advantages, which are key factors in clinical decision-making.

### ROC–AUC Curve for Baseline and Improved ML models

The ROC curves and the respective values of Area Under the Curve (AUC) reported on both the baseline and enhanced machine learning models generate the necessary information on the discriminatory feature of the models in predicting CHD. Figure 19 shows the comparison between four baseline models such as Decision Tree, Random forest, Gradient boosting, and Support Vector Machine (SVM). Of these, the Gradient Boosting and Random Forest models have the highest AUC scores of 0.94, which means that they have a very good chance of separating the CHD–positive and CHD–negative cases. SVM also performs well with an AUC of 0.91, though it lags slightly behind the ensemble models. The Decision Tree model, by contrast, demonstrates the weakest performance, with an AUC of 0.81 and a visibly lower curve, indicating reduced sensitivity and specificity. This underperformance may be attributed to its higher variance and sensitivity to noise in the data.

**Fig 19.**
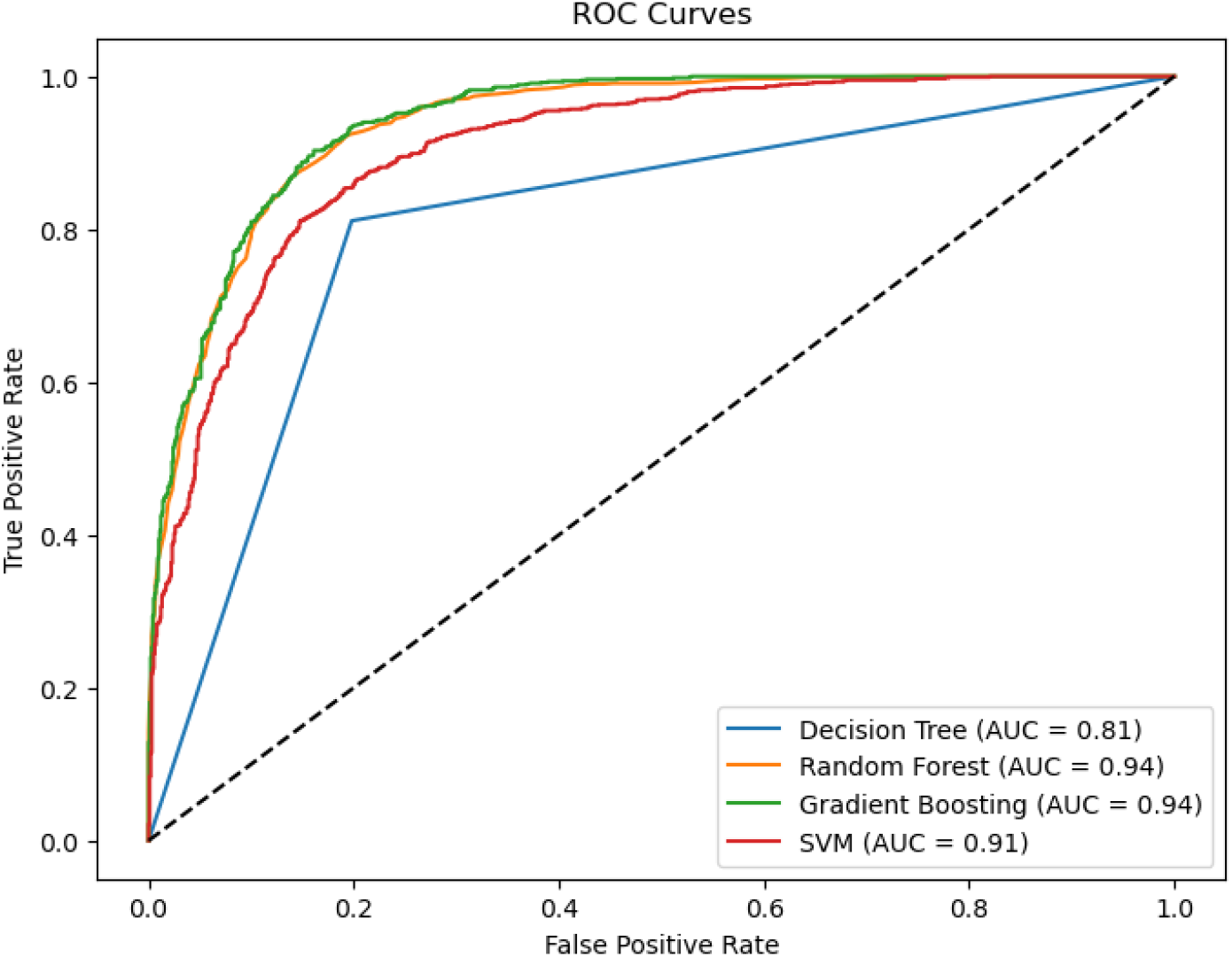
ROC–AUC Curve for Individual (Baseline) ML Models.

Figure 20 illustrates the performance of the improved or enhanced versions of the baseline models—namely, ANRDT (Adaptive Noise-Resistant Decision Tree), HIRF (Hybrid Imbalanced Random Forest), PGBM (Pruned Gradient Boosting Machine), and ESVM (Enhanced Support Vector Machine). The enhancements to the Decision Tree model (ANRDT) led to a remarkable performance boost, with the AUC increasing from 0.81 to 0.93. This suggests that soft splits, pruning, and regularization significantly enhance the model’s robustness and generalization capabilities. Similarly, HIRF achieves an AUC of 0.93, nearly matching the baseline Random Forest, while addressing class imbalance through SMOTE and improving interpretability with SHAP value integration. PGBM retains the high AUC of 0.94 seen in the baseline Gradient Boosting model, indicating that the pruning and outlier resistance techniques did not compromise predictive performance but maintained it while improving model reliability. ESVM, while slightly lower than the baseline SVM with an AUC of 0.90, still demonstrates competitive performance, suggesting that the trade-off for improved interpretability and automated hyperparameter tuning may have come at a minimal cost to classification power.

**Fig 20.**
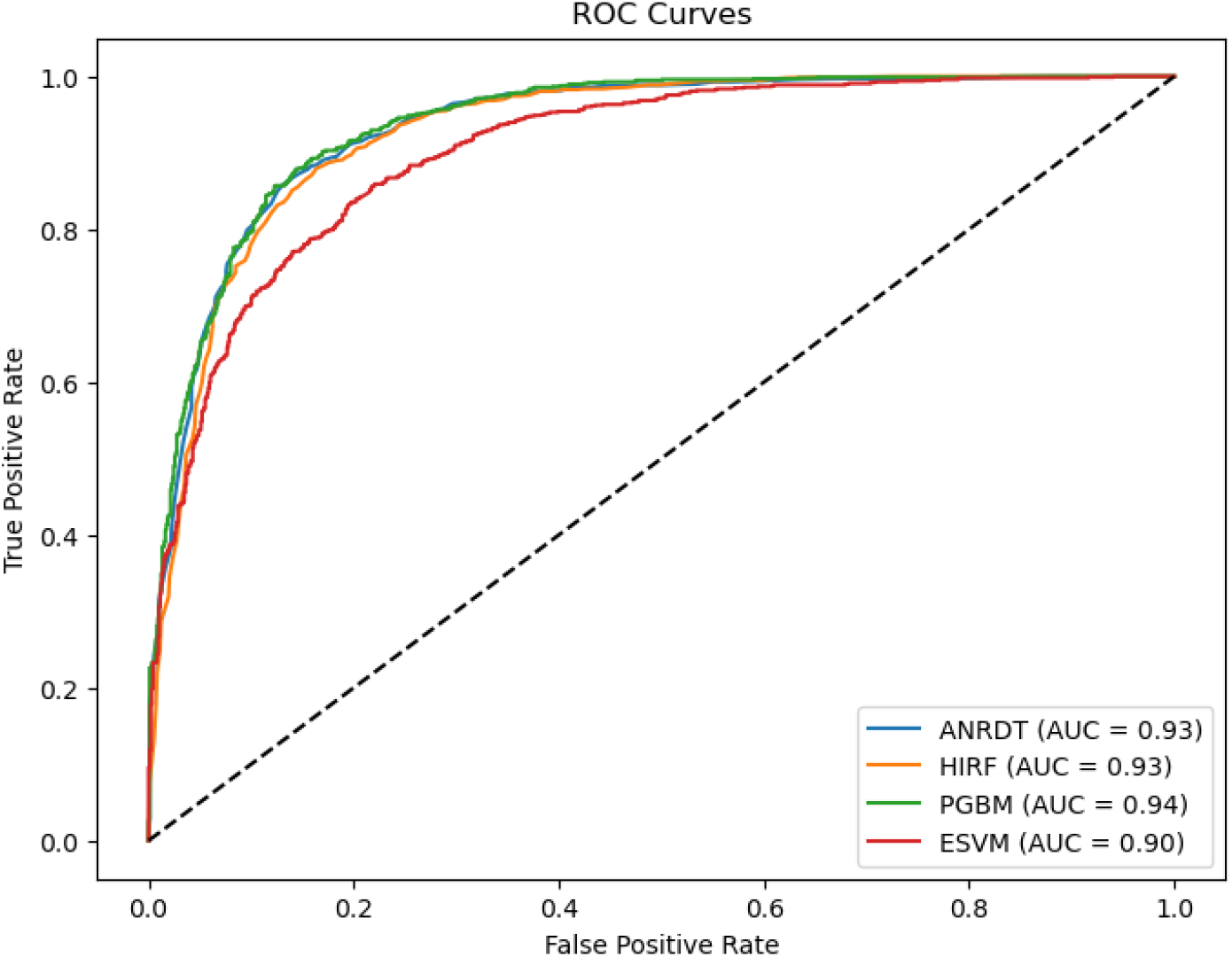
ROC–AUC Curve for Enhanced ML models

A cross–comparison between the two ROC plots reveals that the enhanced models exhibit more tightly grouped and consistently high-performing ROC curves, reflecting their greater robustness and stability across varying thresholds. The improvements in AUC, especially in ANRDT and HIRF, underscore the success of tailored model enhancements in addressing key limitations such as overfitting, class imbalance, and lack of interpretability. These results reinforce the value of hybridization and model optimization strategies in developing machine learning frameworks suited for CHD prediction.

### Precision–Recall Curves

The precision-recall (PR) curves also give a more in-depth analysis of the performance of the machine learning models in predicting CHD, especially when dealing with an imbalanced data set. Figure 21 demonstrates how four basic models: Decision Tree, Random Forest, Gradient Boosting, and Support Vector Machine (SVM) perform. The decision Tree model has the lowest performance among the four, with an average precision (AP) of 0.75. When the recall is higher, its PR curve collapses, and hence, the model is unable to retain its precision when classifying more positive instances. This represents a blatant indication that the Decision Tree produces a relatively large number of false positives, a factor which renders it unsuitable for clinical practice.

**Fig 21.**
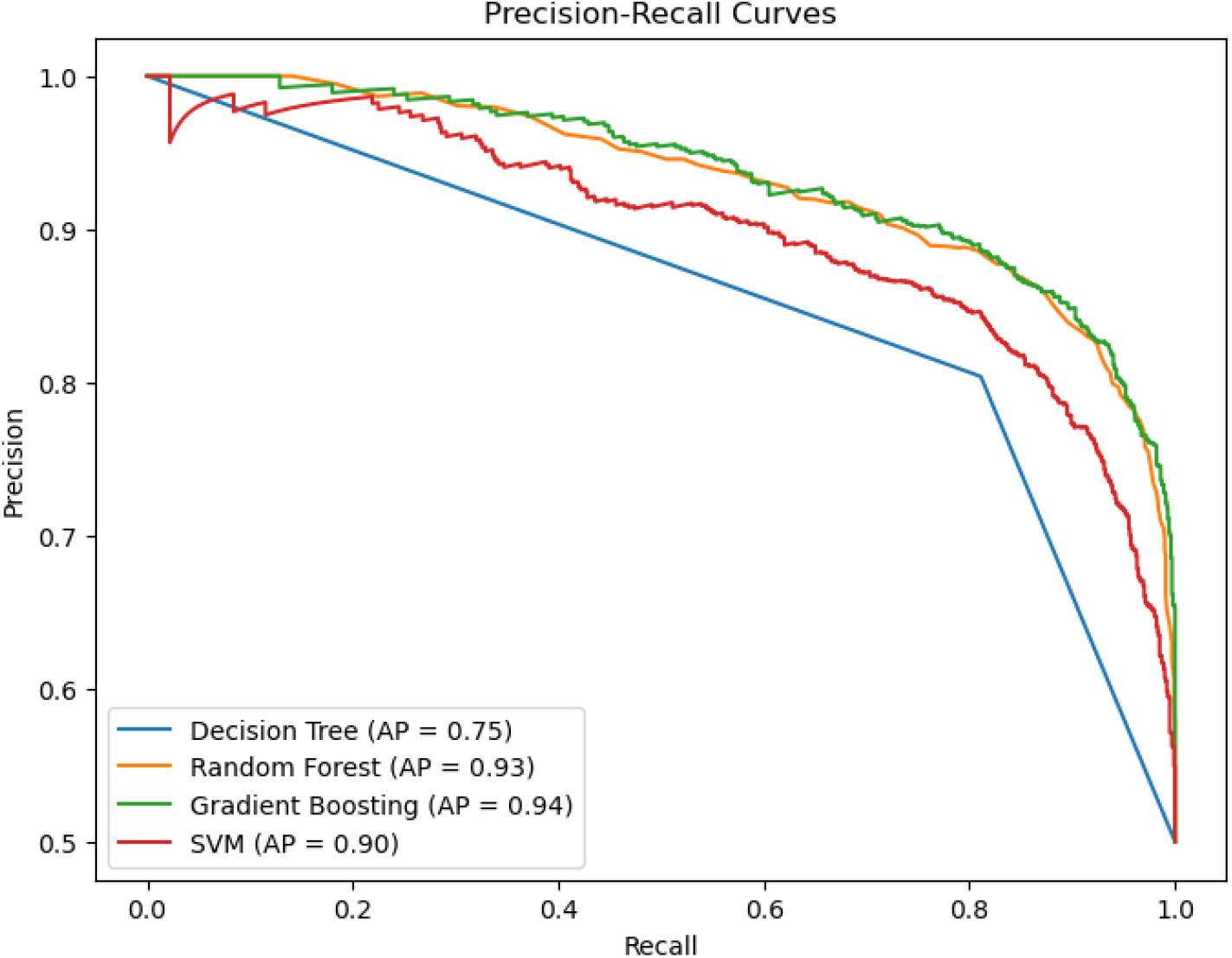
Precision–Recall Curve for Individual (Baseline) ML Models.

By contrast, the Random Forest and Gradient Boosting models perform better, with the highest AP values of 0.93 and 0.94, respectively. Their PR curves are comparatively constant below and near the peak for every level of recall and show a perfect balance of precision and recall. This implies that the two models perform well in the detection of real CHD cases and false positive reduction. The Support Vector Machine (SVM) is also doing a good job as well with an AP of 0.90. Although it is a little below the ensemble models, the SVM has high precision in the majority of the recall thresholds, indicating good and sturdy performance in classification.

To facilitate the investigation of enhanced versions of these models, Figure 22 includes the precision-recall (PR) curves of ANRDT (Adaptive Noise-Resistant Decision Tree), HIRF (Hybrid Imbalanced Random Forest), PGBM (Pruned Gradient Boosting Machine), and ESVM (Enhanced Support Vector Machine). ANRDT excels with a significant change, increasing the AP to 0.92 as compared to the baseline model of 0.75. The positive change highlights the usefulness of noise filtering, pruning, and regularization enhancements to enhance the precision of the Decision Tree and decrease false positives. HIRF also shows a good performance with an AP of 0.91. Though with a slighter decrease than its baseline analogue, the improved model is stable and reliable, particularly when it comes to working with imbalanced data and synthetic oversampling alongside weighted learning.

**Fig 22.**
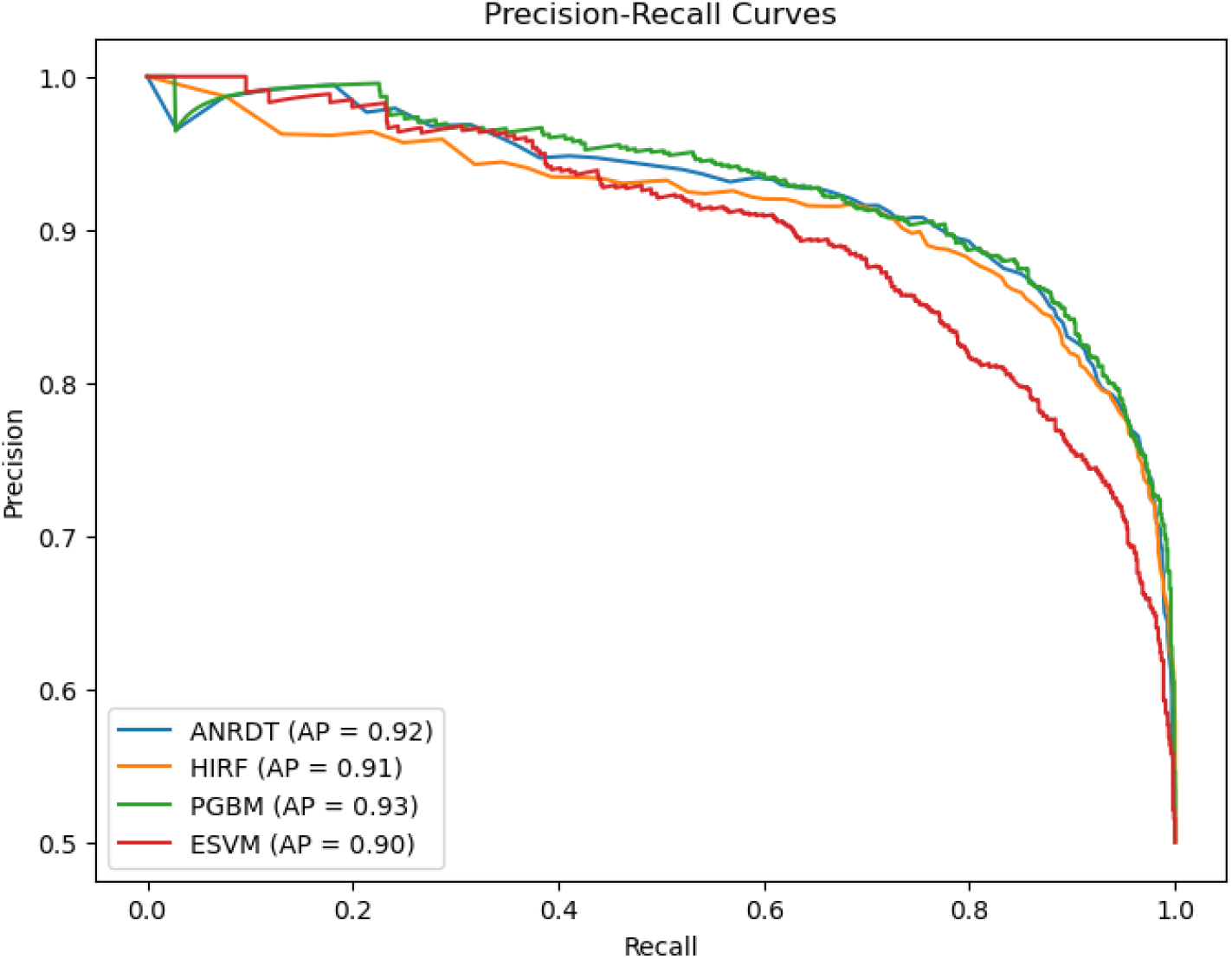
Precision–Recall Curve for Enhanced ML models

PGBM maintains the best performance level with an AP of 0.93, equal to the baseline model Gradient Boosting. The pruning methods and powerful loss functions employed by PGBM enable the model to retain precision and enhance generalizability and interpretability. ESVM is not significantly different with the AP of 0.90 compared to the baseline SVM. The slight dip in average precision may be attributed to the trade–offs introduced by interpretability enhancements and automated tuning, yet the curve still indicates stable and reliable classification performance.

### Calibration Curves for Baseline and Enhanced ML Models

The calibration curves shown in Figures 23 and 24 provide critical insights into the reliability of predicted probabilities for CHD. Calibration assesses how closely the predicted probabilities from a model reflect the actual likelihood of the outcome. Ideally, a model’s calibration curve aligns with the diagonal 45–degree line, indicating perfect correspondence between predicted and observed probabilities.

**Fig 23.**
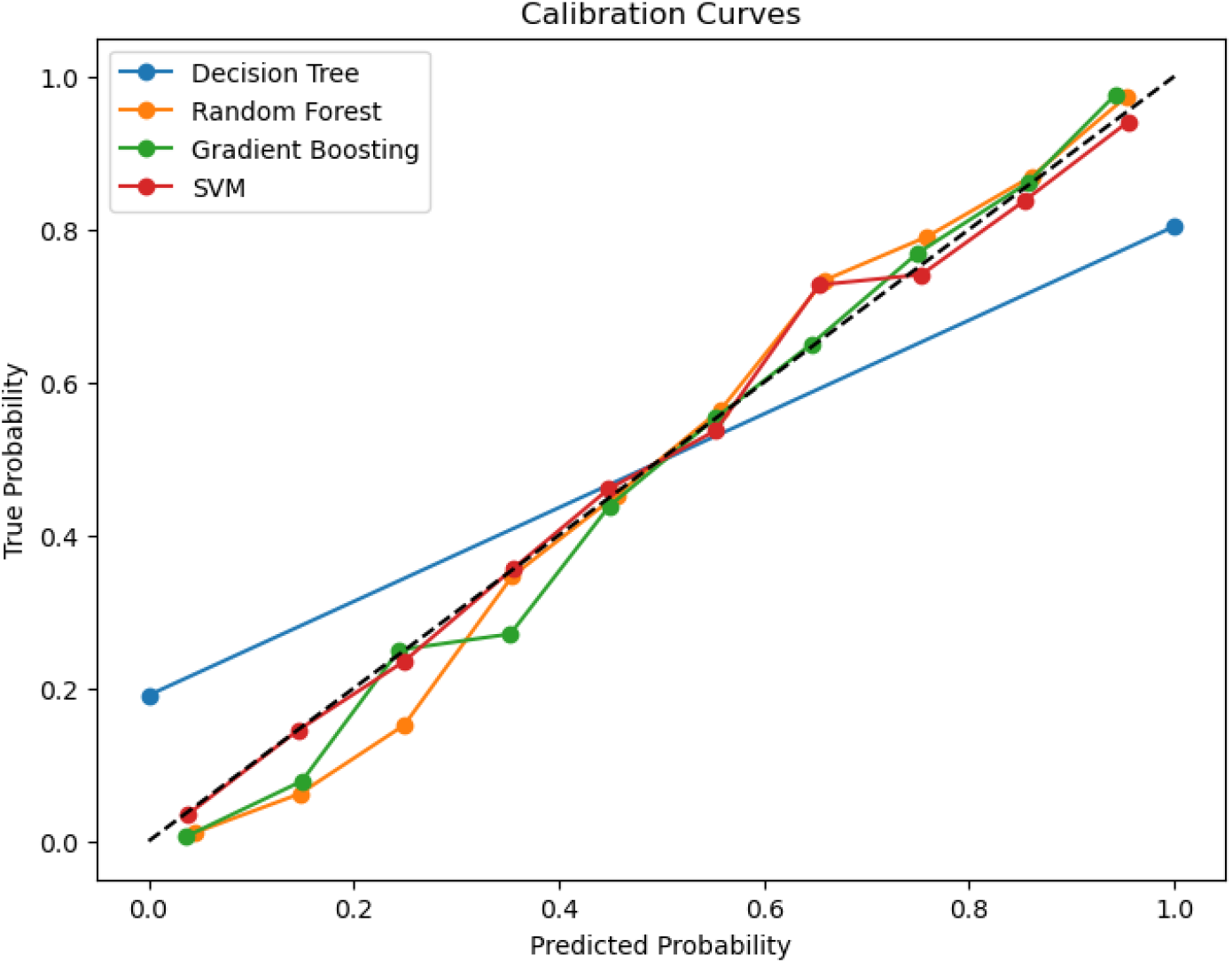
Calibration Curve for Baseline ML Models

**Fig 24.**
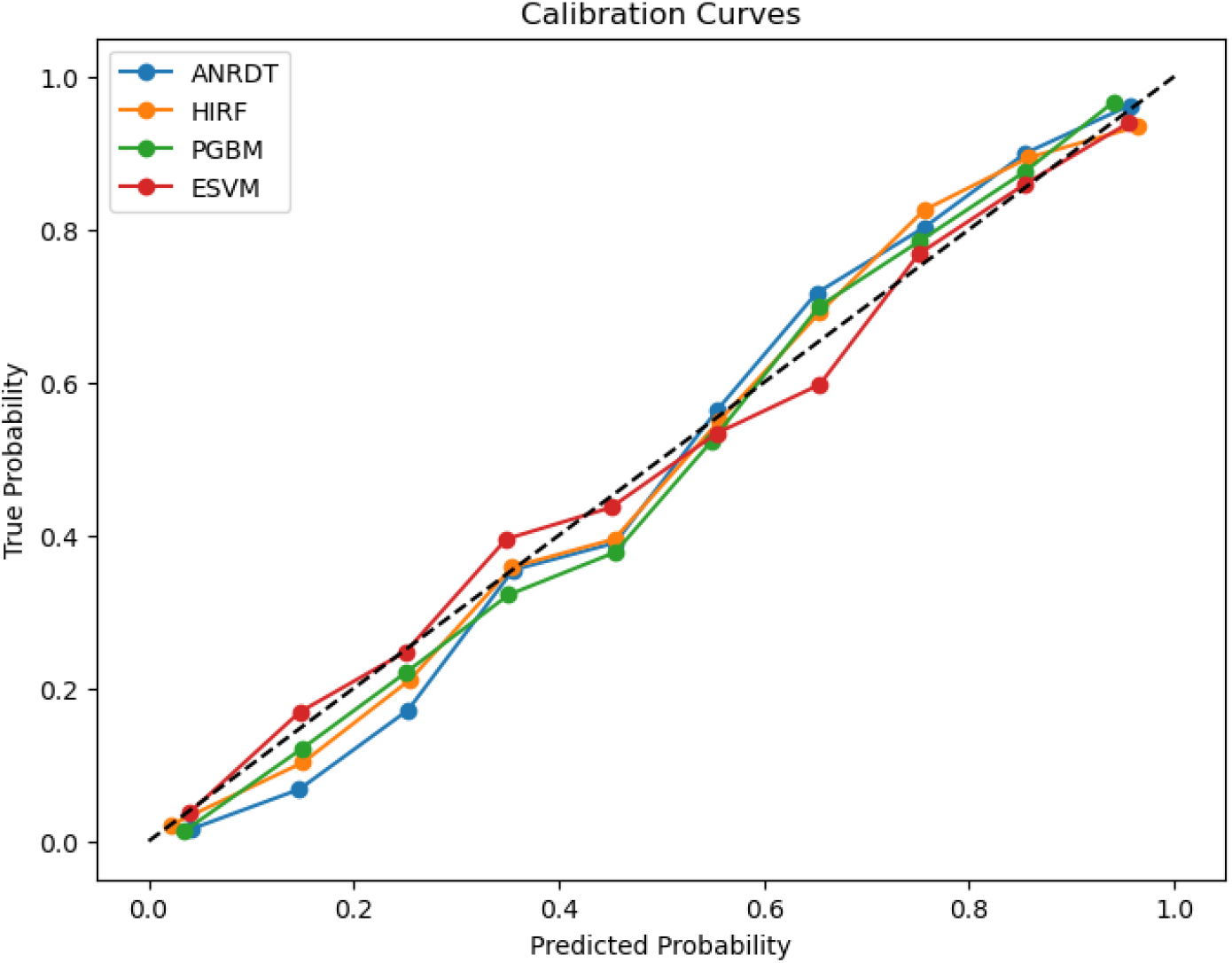
Calibration Curve for Enhanced ML Models

In Figure 23, the calibration performance of the baseline models–Decision Tree (DT), Random Forest (RF), Gradient Boosting (GBM), and Support Vector Machine (SVM)–is illustrated. The Decision Tree exhibits notable miscalibration, with its curve consistently deviating above the diagonal line. This indicates overconfidence in its predictions, where predicted probabilities are higher than actual observed outcomes.

This is consistent with the model’s relatively lower performance across accuracy and AUC–ROC metrics. In contrast, both Random Forest and Gradient Boosting maintain calibration curves closer to the diagonal, particularly at mid to high probability ranges, suggesting reliable probability estimates. The SVM model demonstrates moderate calibration, with a slight tendency to underestimate probabilities, particularly in the mid-range. This conservative calibration is characteristic of SVMs and contributes to their generally stable but sometimes less sensitive classification behavior.

Figure 24 shows the calibration curves of the enhanced models Adaptive Noise-Resistant Decision Tree (ANRDT), Hybrid Imbalanced Random Forest (HIRF), Pruned Gradient Boosting Machine (PGBM), and Enhanced Support Vector Machine (ESVM). These models are better calibrated than their baseline counterparts. ANRDT shows substantial improvement upon the contrast of baseline DT, with its calibration curve being a far better fit with the ideal diagonal. This symbolizes the optimistic effect of reinforcements like probabilistic splitting, noise filtering and regularization in alleviating overfitting and enhancing the reliability of predictions. HIRF and PGBM are also well-calibrated and highly coherent with the diagonal over the range of probabilities. These enhancements align with their good classification responsiveness and add that the algorithmic improvement is not only accuracy improvements but also confidence estimation. ESVM calibration profile is close to the baseline SVM with marginal underestimation of probabilities despite its overall reliable performance.

### Learning Curves for Baseline VS. Enhanced ML Models Baseline ML Models

The learning curve for the Decision Tree (DT) model is presented in Figure 25. The training score remains at a perfect 1.00 across all training set sizes, indicating that the model fully memorizes the training data. In contrast, the cross-validation (CV) score starts relatively low, at around 0.75, and increases modestly to approximately 0.78.

**Fig 25.**
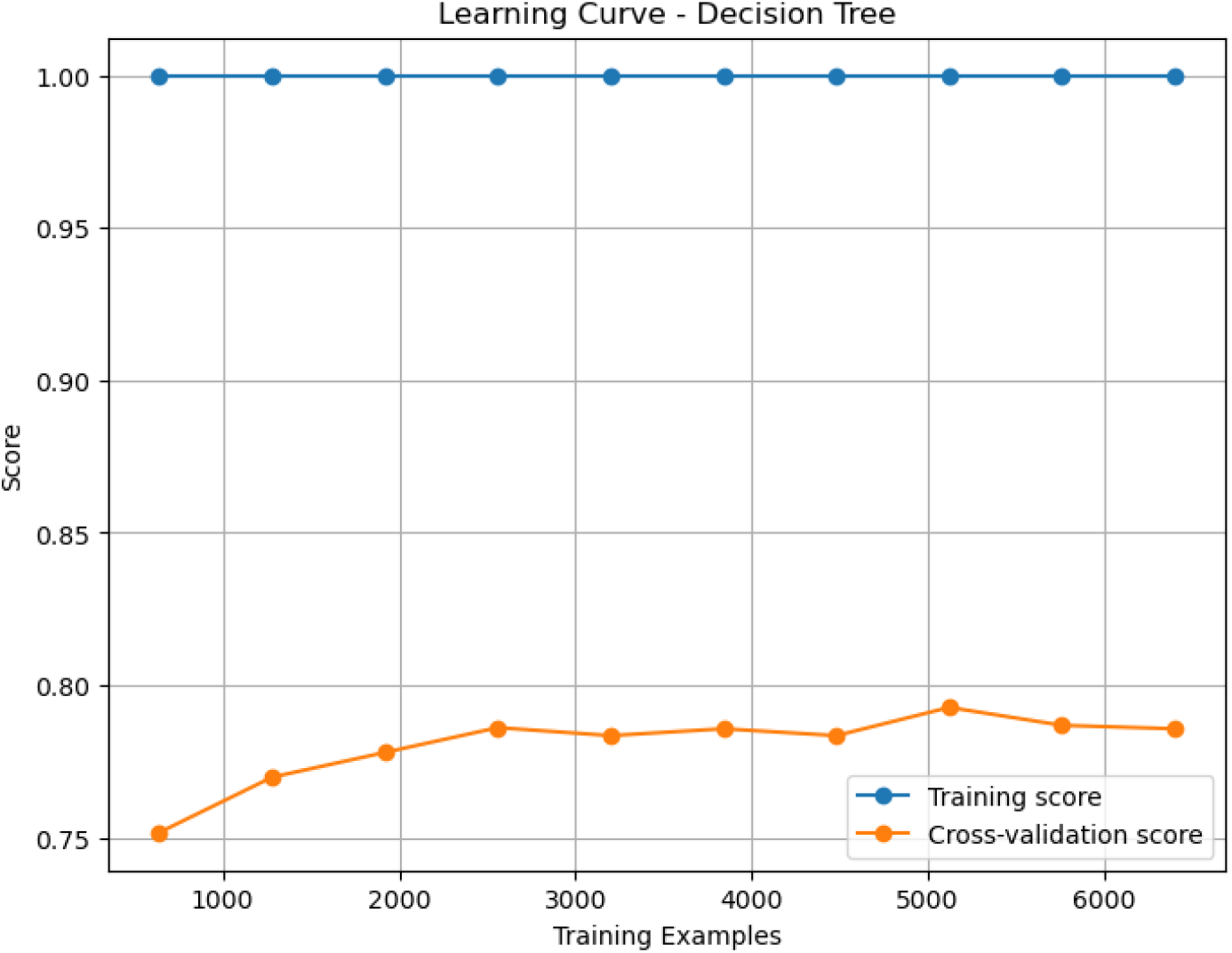
Learning Curve for Decision Tree

Despite this slight improvement, a substantial gap persists between the training and validation scores, clearly demonstrating overfitting. The DT model captures intricate patterns in the training data but fails to generalize to unseen data. This behavior is typical of unpruned or overly complex decision trees, which are prone to learning noise, resulting in high variance. The limited gain in validation performance with increasing data suggests that additional training samples alone are insufficient to resolve overfitting. Instead, techniques such as pruning, regularization, or switching to ensemble approaches may be necessary to enhance generalization.

Figure 26 shows the learning curve for the Random Forest (RF) model. Similar to DT, the training score remains close to 1.00, reflecting the model’s capacity to fit the training set well. However, the CV score starts higher than in DT, around 0.84, and improves gradually to about 0.86. While there is still a gap between training and validation scores, it is narrower compared to DT, indicating reduced overfitting. This improvement is attributed to the ensemble nature of RF, which leverages bagging and random feature selection to reduce variance. Although some overfitting remains, the model generalizes better than DT, making RF a more reliable baseline model. The plateauing of the CV score suggests that further enhancements may require hyperparameter tuning (e.g., number of estimators, tree depth) or additional feature engineering.

**Fig 26.**
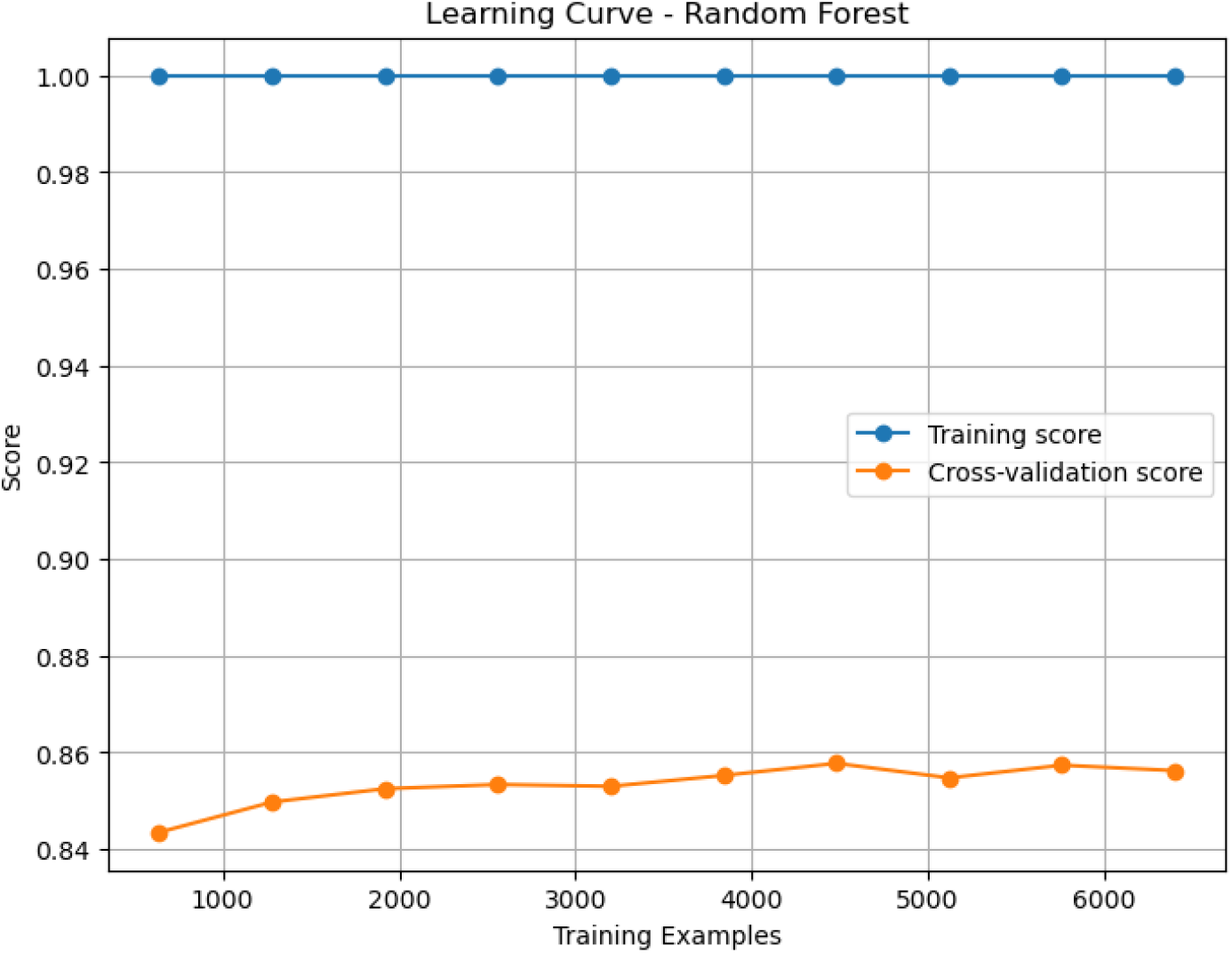
Learning Curve for Random Forest

The learning curve for the Gradient Boosting Machine (GBM) model is presented in Figure 27. The training score starts high at around 0.945 and gradually declines to just below 0.88 as more training data is added. This downward trend indicates that the model becomes less dependent on specific patterns in the training data and starts learning more generalizable relationships. In parallel, the CV score begins at approximately 0.845 and steadily increases, plateauing near 0.861. The moderate and decreasing gap between training and validation scores suggests that GBM manages overfitting more effectively than DT or RF. Unlike RF, GBM builds trees sequentially, with each new learner correcting the errors of the previous ones. This sequential learning process enhances accuracy but can also introduce sensitivity to noise. The stable CV score after a certain data threshold implies that the model has captured most of the useful patterns in the data. Further improvement would likely depend on regularization strategies such as shrinkage, subsampling, or adjusting the number of boosting iterations and learning rate.

**Fig 27.**
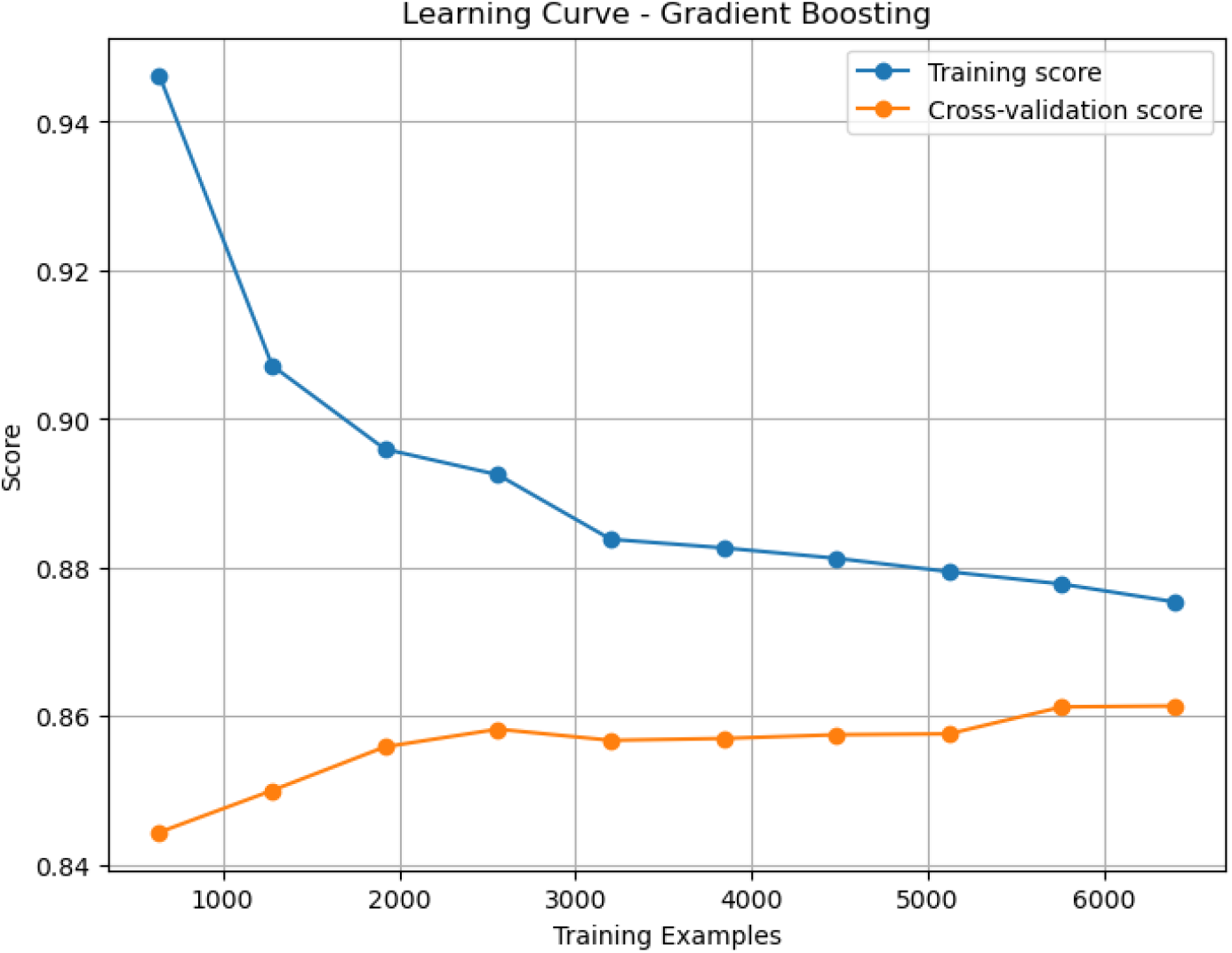
Learning Curve for Gradient Boosting Machine (GBM)

Figure 28 illustrates the learning curve for the Support Vector Machine (SVM) model. Both training and CV scores begin relatively low—around 0.66 and 0.63, respectively—but exhibit a consistent upward trend with increasing training size. The training score gradually rises to approximately 0.825, while the CV score closely follows, ending around 0.82. The consistently narrow gap between the two curves indicates low variance and strong generalization. This is characteristic of SVMs, which, when properly regularized and used with suitable kernels, effectively manage the bias-variance tradeoff. Unlike tree-based models, which tend to overfit, SVMs aim to maximize the margin between classes, which helps prevent overfitting, particularly in high-dimensional or noisy datasets. The steady improvement in CV performance suggests that the SVM model benefits significantly from additional training data and has not yet exhausted its learning capacity.

**Fig 28.**
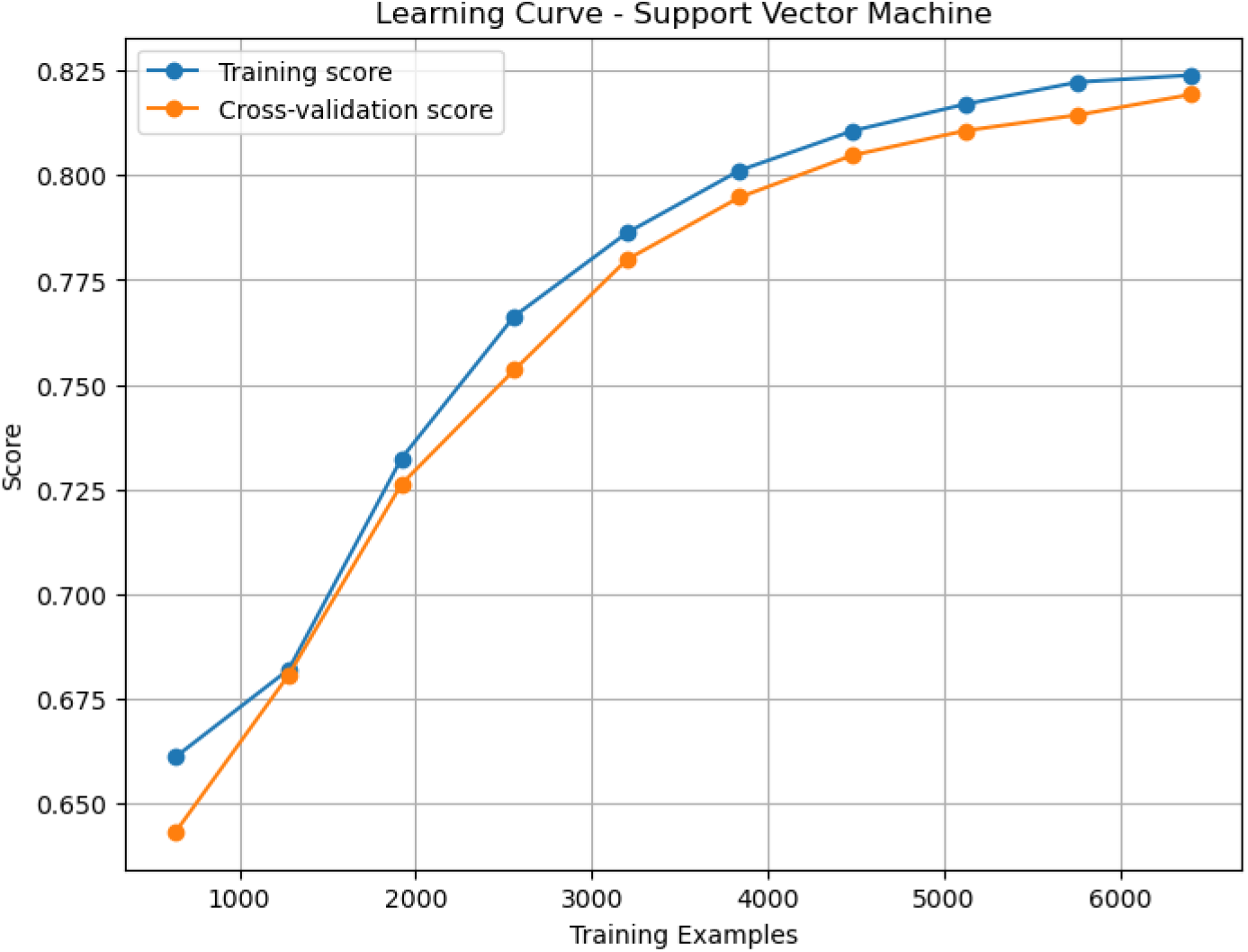
Learning Curve for Support Vector Machine (SVM)

### Enhanced ML Models

Learning curves are valuable diagnostic tools for understanding a model’s performance as the size of the training dataset increases. They display two key metrics: the training score (performance on the training set) and the cross-validation (CV) score (performance on unseen data), both plotted against training set size. A large, persistent gap between these curves indicates overfitting, while convergence suggests effective generalization.

The learning curve for the Adaptive Noise-Resistant Decision Tree (ANRDT), illustrated in Figure 29, shows that the training score remains consistently perfect at 1.00 across all training set sizes. This reflects the model’s ability to fully memorize the training data. However, the CV score starts at approximately 0.845 and rises only slightly to around 0.857. The wide and unchanging gap between the training and CV scores clearly indicates a high degree of overfitting, suggesting that ANRDT struggles to generalize to new data. Although there is a minor improvement in CV performance with more data, it is insufficient to offset the overfitting. To improve generalization, strategies such as regularization or pruning would be necessary.

**Fig 29.**
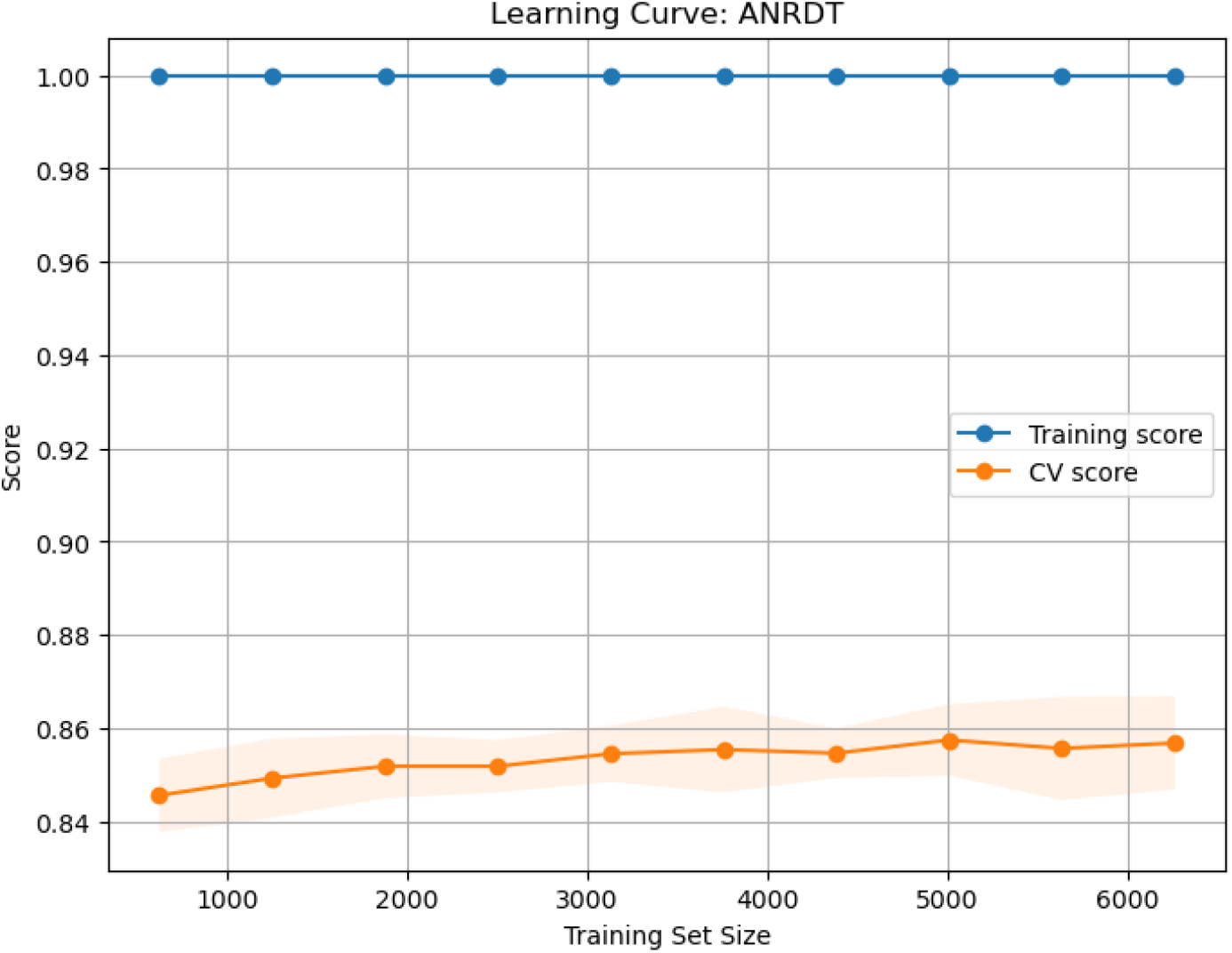
Learning Curve for ANRDT

Figure 30 presents the learning curve for the Hybrid Imbalanced Random Forest (HIRF). Similar to ANRDT, the training score remains at 1.00 across all training sizes, indicating strong memorization of the training set. However, the CV score starts at around 0.83 and increases more steadily, reaching approximately 0.853. While overfitting is still present, it is less pronounced than in ANRDT. The narrowing gap between the training and CV scores suggests that HIRF benefits more effectively from increasing training data, likely due to its ensemble nature and mechanisms for addressing class imbalance. These properties enhance its generalization performance compared to ANRDT.

**Fig 30.**
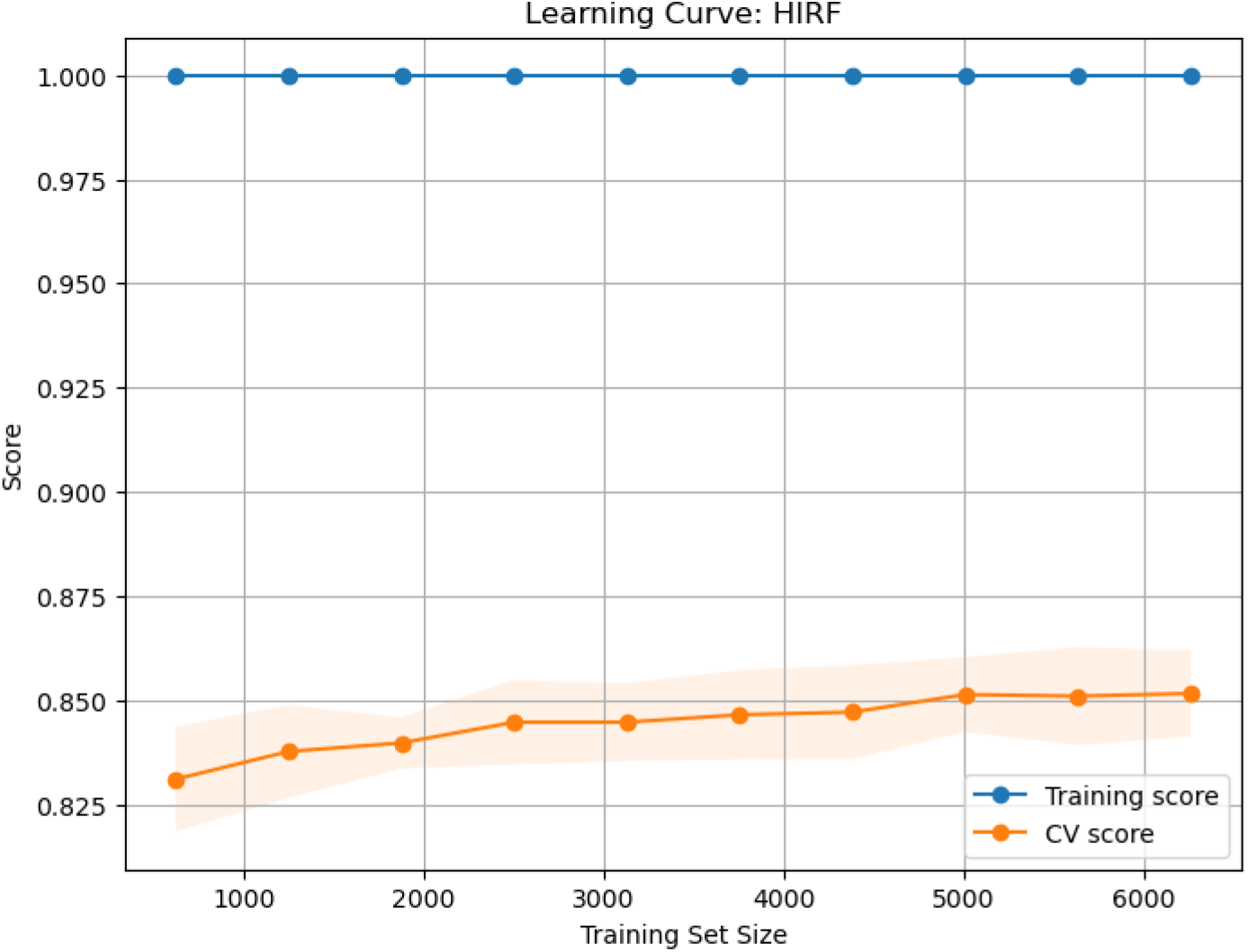
Learning Curve for HIRF

On the contrary, the Pruned Gradient Boosting Machine (PGBM) learning curve in Figure 31 illustrates a more balanced learning pattern. The training score begins with an initial high score of around 0.96, lowering slowly with an increase in the size of training to around 0.88. Meanwhile, the CV score level increases systematically by approximately 0.84 to 0.864. The overlap of training scores and CV scores shows that the bias-variance tradeoff is in reasonable control. This means that the pruning techniques used in PGBM help counteract overfitting. The PGBM model is one of the improved models that have a good tradeoff between accuracy and generalizability, which makes it deployable in CHD risk prediction in the real world. Figure 32 presents the learning curve of the Enhanced Support Vector Machine (ESVM). At the initial stages, training and CV scores are similarly low, commencing at around 0.66 and 0.60, respectively. Nonetheless, both measures show a steady and corresponding increase in terms of the size of the training set. In the end, they meet at the 0.83 value. This trend represents excellent generalizability and low overfitting, meaning that the ESVM is capable of learning with additional data and achieving more reliable predictions. This kind of behavior means that ESVM is a promising choice for scalable, reliable CHD prediction, especially where interpretability and consistency take precedence.

**Fig 31.**
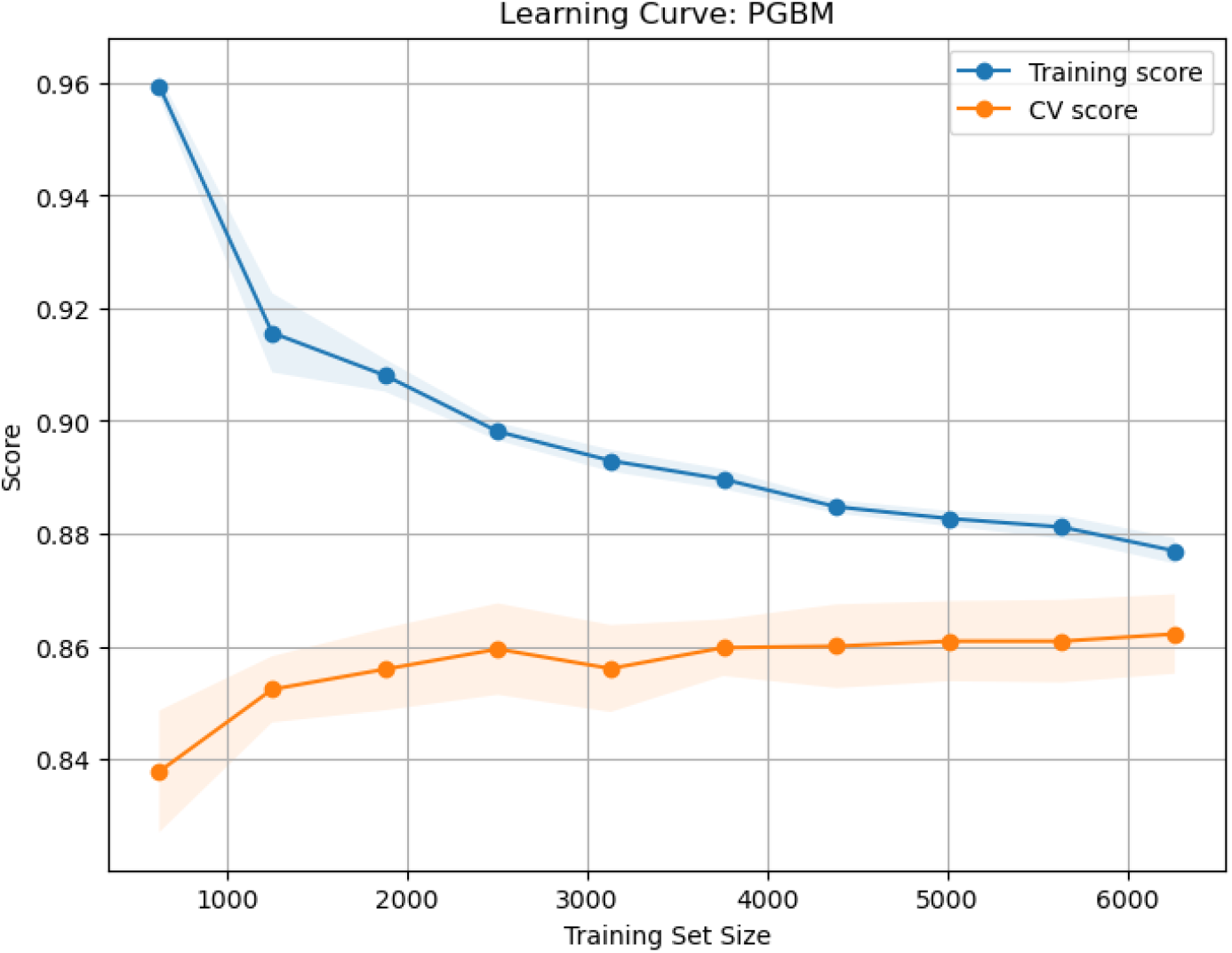
Learning Curve for PGBM

**Fig 32.**
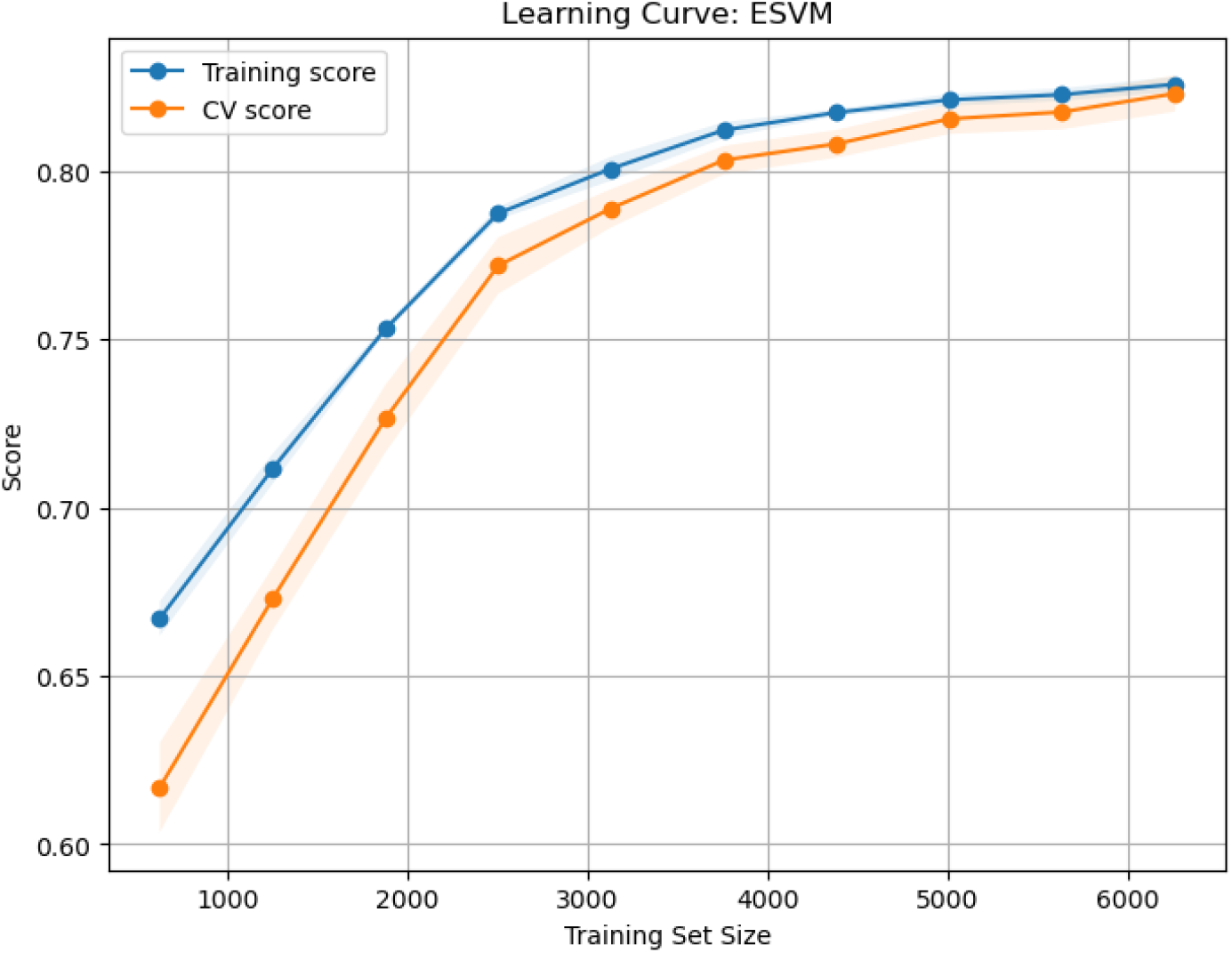
Learning Curve for ESVM

### Comparison of Learning Curves: Baseline vs. Enhanced Models

The juxtaposition of the learning curves of the baseline and the improved machine learning models gives a critical insight into the generalization behavior of the models and the ability to overcome overfitting. In the baseline models, the Decision Tree (DT) performs excellently on the training set with all sample sizes, implying that it is overfitting the data. Nevertheless, its cross-validation (CV) score is much lower, and it has small incremental training progress. The tendency of the training performance to remain high but the validation performance to remain lower is a clear sign of overfitting, and it is indicative that the model fits the noise and certain patterns specific to the training data and fails to generalize to the validation set. The Random Forest (RF) model also does not overfit quite as much as the Decision Tree (DT), with the gap between the training and the validation scores being much narrower. This can be explained by its ensemble architecture that integrates variance-reduction schemes, which are bootstrapping and random feature selection. However, the plateau in the validation score implies that the potential of RF will not be high until it is tuned or artifacts are added to the data.

Gradient Boosting Machine (GBM) has a better learning dynamic. With an increase in the size of the training set, the training accuracy of GBM is minimally reduced, whereas the CV score is consistently increased. The trend indicates the ability of the model to learn sequentially by drawing on previous errors and thus generalizing better than DT and RF. Nevertheless, the final difference between the training and validation scores suggests that GBM will be prone to overfitting unless adequately regularized.

The Support Vector Machine (SVM) indicates the most balanced performance among the models that form the baseline analysis. Its learning curves, training, and validation have relatively low starting but maintain the same monotonic growth pattern. The decrease in the distance between them indicates a high level of generalization and indicates that SVM heuristically manages the bias-variance problem well, at least when used with a proper kernel and regularisation strategy.

On the contrary, the improved models have better generalization behavior and improved learning when additional training data is provided. The Adaptive Noise-Resistant Decision Tree (ANRDT) reflects the overfitting pattern of the baseline DT. Accordingly, the Adaptive Noise-Resistant Decision Tree also reached a perfect training score, with cross-validation (CV) performance being relatively flat. Although designed to be robust against noise, ANRDT generalizes poorly, suggesting that it might be necessary to augment it with some further mechanism, e.g., pruning or regularisation, before it can convert its memorization ability into prediction accuracy. Nevertheless, the Hybrid Imbalanced Random Forest (HIRF) demonstrates a more promising dynamic. Although it continues to perform well in terms of training accuracy, its validation performance improves more consistently as compared to ANRDT. This implies that its improvements in dealing with imbalanced data, possibly as a result of weighting or superior sampling methods, will be a contributor to improved generalization.

An exciting improvement is noted in the Pruned Gradient Boosting Machine (PGBM), displaying a favorable balance of bias-variance tradeoff. As the training set size rises, the accuracy of the model in training gradually drops, but the cross-validation score constantly improves and then reaches a plateau slightly below the training curve. This overlap shows efficient pruning and regularization that enables PGBM to attain high accuracy without overfitting. The comparison of PGBM and its baseline version shows a more stable and generalizable result. Looking at it the same way, Enhanced Support Vector Machine (ESVM) also has a good generalization ability. After starting both training and validation scores at low levels, the difference in the two scores is seen to decrease with an increase in data, indicating a close convergence. This trend suggests that ESVM is capable of rectifying the underlying configuration of the data rather than being subjected to noise, probably because of better regularization and a better selection of kernels.

Generally, the improved models are shown to enhance the learning efficiency and the generalization ability of the model over the baseline models. Although techniques to improve ensembles, such as HIRF, have some effect in discouraging overfitting, much greater improvements can be achieved with more focused modifications, such as pruning, boosting optimization, and regularisation, which are observed in models that place a stronger emphasis on model customization, such as PGBM and ESVM. Such findings highlight the importance of complexity control and robustness strategies in model improvement as a means of constructing solid and scalable predictive systems in medicine, fields like predicting coronary heart disease risk.

### Ensemble Learning Models

The performance comparison between baseline and enhanced ensemble learning models reveals important patterns in how each approach contributes to coronary heart disease (CHD) prediction. All ensemble methods exhibit high predictive capacity, with accuracies generally above 0.85. Among the baseline models, stacking achieves the highest overall accuracy at 0.872, followed closely by boosting at 0.866. Stacking also demonstrates a strong balance across precision (0.854), sensitivity (0.898), and specificity (0.846), indicating its robustness in classifying both positive and negative CHD cases. Boosting, although slightly lower in precision and specificity, yields the highest sensitivity (0.908), highlighting its strength in correctly identifying positive CHD cases, albeit at a slight cost to false-positive rates.

Majority voting also performs competitively, with an accuracy of 0.864, as well as well-balanced precision (0.857) and specificity (0.854). Its relatively simple decision mechanism makes it effective for stabilizing predictions across diverse base learners. Bayesian Model Averaging (BMA), though theoretically powerful due to its probabilistic weighting of model outputs, delivers slightly lower performance (accuracy of 0.858), possibly due to challenges in estimating accurate posterior probabilities with limited data. Bagging, on the other hand, while slightly underperforming in precision and specificity compared to other methods, offers the second-highest sensitivity (0.882), making it valuable when minimizing false negatives is critical.

The enhanced ensemble models, which are built by combining optimized versions of individual base learners, show marginal but meaningful improvements or stability in performance. The enhanced stacking model maintains strong performance across all metrics, achieving an accuracy of 0.868, a precision of 0.852, and balanced recall and specificity of 0.891 and 0.845, respectively. This suggests that integrating improved base learners helps retain stacking’s advantage in generalization. Enhanced boosting also sustains its high sensitivity (0.891), indicating its continued effectiveness in recognizing CHD cases after enhancement, while its specificity (0.842) improves compared to the baseline. Similarly, enhanced majority voting maintains its original accuracy (0.862) and shows a slight improvement in precision, reinforcing the utility of simple ensemble strategies when paired with stronger individual classifiers.

Interestingly, enhanced BMA sees a slight uptick in accuracy (from 0.858 to 0.862) and sensitivity (from 0.876 to 0.886), suggesting that when better-calibrated models are used as inputs, BMA’s probabilistic framework becomes more effective. Enhanced bagging, however, maintains a similar performance profile to its baseline counterpart, with nearly identical metrics. This implies that the improvements in individual learners may not substantially alter bagging’s inherent variance-reduction mechanism unless its architecture is further modified (e.g., through feature selection or deeper trees).

Table 9 presents the detailed classification metrics for both baseline and enhanced ensemble models.

**Table 9.** Comparison of baseline and enhanced ensemble models on classification metrics.

| Model | Accuracy | Precision | Sensitivity | Specificity |
| --- | --- | --- | --- | --- |
| *Baseline Ensemble Models* |  |  |  |  |
| Bayesian Model Averaging (BMA) | .858 | .845 | .876 | .839 |
| Majority Voting | .864 | .857 | .873 | .854 |
| Bagging | .856 | .838 | .882 | .830 |
| Boosting | .866 | .838 | .908 | .824 |
| Stacking | .872 | .854 | .898 | .846 |
| *Enhanced Ensemble Models* |  |  |  |  |
| BMA (Enhanced) | .862 | .846 | .886 | .839 |
| Majority Voting (Enhanced) | .862 | .856 | .870 | .854 |
| Bagging (Enhanced) | .856 | .838 | .884 | .829 |
| Boosting (Enhanced) | .866 | .849 | .891 | .842 |
| Stacking (Enhanced) | .868 | .852 | .891 | .845 |

### Comparative Evaluation of Machine Learning and Ensemble Models for CHD Prediction

The comparative evaluation of ML models, and ensemble learning models for predicting CHD reveals critical insights into the strengths and trade-offs of each modelling approach.

### Individual (Baseline) Machine Learning Models

Among the baseline machine learning models, Random Forest, Gradient Boosting, and Support Vector Machine (SVM) achieved competitive results. Gradient Boosting reached an accuracy of 86.6%, with high sensitivity (90.8%) but slightly lower precision (83.8%) and specificity (82.4%). Random Forest followed closely with an accuracy of 86.3% and more balanced metrics. SVM, while somewhat less accurate (83.0%), offered reasonably high sensitivity (84.9%) and precision (81.7%), demonstrating its utility in balanced classification tasks.

Enhanced versions of these models showed moderate but meaningful improvements. The Pruned Gradient Boosting Machine (PGBM), for example, preserved the original model’s high performance with improved precision (84.9%) and specificity (84.2%). The Adaptive Noise-Resistant Decision Tree (ANRDT) significantly enhanced baseline decision trees, achieving an accuracy of 86.2%–approaching ensemble-level performance. These findings highlight the potential of model-specific enhancements to close the performance gap with more complex ensembles while retaining interpretability and efficiency.

### Ensemble Learning Models

The current results demonstrate that ensemble learning models succeed in predicting CHD more effectively than classical statistical models and even single machine learning algorithms. These models exploit the phenomenon of merging the different learners-each with their predictive advantages, to produce a higher and better overall model.

Stacking was the best-performing model of all tested models. The baseline stacking model performed with an accuracy of 87.2%, precision of 85.4%, sensitivity of 89.8%, and specificity of 84.6%, which portrays a high overall balance, and there was little trade–off between false positive and false negative. These measures indicate that not only is stacking precise but also clinically valid, especially in medical uses where over and under-diagnosis is crucial. The improved stacking model retained this performance advantage, with an accuracy of 86.8% and a slightly modified sensitivity of 89.1%, which showed the stability of the method despite tuning and feature optimization.

The other crucial ensemble technique, boosting, did incredibly well on both its default and improved versions as well. Gradient Boosting Machines (GBM) in the ensemble framework yielded the highest sensitivity of 90.8%, among all the tested models. The resulting high sensitivity is essential in CHD–screening and triage in which any failure to detect high–risk individuals would result in dire consequences–an advantage of boosting is that it iteratively refines the errors of the previous model, enabling the ensemble to target the more difficult-to-classify cases–an advantage in the case of imbalanced datasets, such as BRFSS. With enhanced boosting (PGBM), the accuracy was unchanged (86.6%), and precision and specificity were slightly increased, demonstrating that boosting algorithms can benefit from preprocessing steps like pruning and class weighting. Boosting can be an important choice as a standalone tool or in larger predictive pipelines, as it is highly reliable and also highly scalable.

Other ensemble methods, including Bayesian Model Averaging (BMA), Majority Voting, and Bagging, performed reliably and steadily as well. BMA obtained an accuracy of 85.8% for the baseline model and 86.2% for the best model, having similar precisions and specificities, demonstrating stability. Majority Voting also has a comparable precision of over 85% and sensitivity near 87% as well, thus being a viable alternative when simplicity and explainability are required. Bagging models are less precise and specific than other ensembles, but they have high sensitivity with both the baseline (88.2%) and the enhanced form (88.5%), so they are appropriate when it is most important to identify real positive cases. These results demonstrate that even primitive ensembles can dramatically outperform individual learners through their ability to decrease variance and balance a model bias, which is of special importance in clinical data where the variables may be noisy or correlating.

The high performance of the ensemble models can be attributed to a few important benefits. One is that they minimize the problem of overfitting by averaging the decisions of many different learners, which regularizes extreme predictions and reduces outlier impacts. Second, they increase generalization on unseen data, which can be proven by their stability in cross-validation folds in this evaluation. Third, ensemble approaches are flexible and can, therefore, allow the use of both interpretable models (e.g., logistic regression) and high-scoring black-box learners (e.g., GBM, SVM), which are useful in a hybrid clinical environment. Enhancement of base learners used in this study, like Adaptive Noise–Resistant Decision Tree (ANRDT) and Pruned Gradient Boosting Machine (PGBM), provides an additional advantage in terms of enhancing ensemble reliability by alleviating noise sensitivity and effectively managing class imbalances. The consistency of ensemble models in leading all the main metrics, including accuracy, precision, sensitivity, and specificity, demonstrates their feasibility of clinically applicable application.

The other advantage of ensemble learning is that it can adjust to real-world constraints. Computational resources are usually scarce in LMICs such as Kenya, and the interpretability of models is pivotal toward adoption by non-technical healthcare workers. Ensemble methods like stacking built using interpretable base learners are a trade-off between transparency and predictive performance. Explainability in this study was facilitated by the SHAP (Shapley Additive Explanations), which enabled us to understand which features had the most impact on CHD predictions given a set of ensemble learners. This will allow de-mystifying complex predictions and proving clinical trust in AI-driven tools. Moreover, the high calibration rates of the ensemble models make their probability results sound enough to be incorporated in risk communication to patients or be part of automated decision-support processes, which constitutes a critical feature in health applications.

A detailed summary of model performance across all evaluated methods—including classical, baseline machine learning (ML), and enhanced ensemble models—is presented in Table 10, illustrating the comparative strengths and trade-offs of each approach.

**Table 10.** Comparison of individual machine learning and ensemble models on CHD prediction metrics.

| Model | Accuracy | Precision | Sensitivity | Specificity |
| --- | --- | --- | --- | --- |
| *Baseline Machine Learning Models* |  |  |  |  |
| Decision Tree | .807 | .804 | .811 | .802 |
| Random Forest | .863 | .845 | .888 | .837 |
| Gradient Boosting | .866 | .838 | .908 | .824 |
| Support Vector Machine | .830 | .817 | .849 | .810 |
| *Baseline Ensemble Models* |  |  |  |  |
| Bayesian Model Averaging (BMA) | .858 | .845 | .876 | .839 |
| Majority Voting | .864 | .857 | .873 | .854 |
| Bagging | .856 | .838 | .882 | .830 |
| Boosting | .866 | .838 | .908 | .824 |
| Stacking | .872 | .854 | .898 | .846 |
| *Enhanced Machine Learning Models* |  |  |  |  |
| ANRDT (Enhanced DT) | .862 | .848 | .883 | .842 |
| HIRF (Enhanced RF) | .856 | .838 | .884 | .829 |
| PGBM (Enhanced GBM) | .866 | .849 | .891 | .842 |
| ESVM (Enhanced SVM) | .816 | .790 | .861 | .771 |
| *Enhanced Ensemble Models* |  |  |  |  |
| BMA (Enhanced) | .862 | .846 | .886 | .839 |
| Majority Voting (Enhanced) | .862 | .856 | .870 | .854 |
| Bagging (Enhanced) | .856 | .838 | .884 | .829 |
| Boosting (Enhanced) | .866 | .849 | .891 | .842 |
| Stacking (Enhanced) | .868 | .852 | .891 | .845 |

## Discussion

The results of this study demonstrate that enhanced ML models and hybrid ensemble strategies offer significant improvements in predicting CHD, particularly when compared to traditional statistical methods. While classical models such as logistic regression and Cox proportional hazards are valued for their interpretability, they are often constrained by assumptions of linearity and limited capacity to capture interactions among predictors [24], [21]. In contrast, our findings indicate that machine learning (ML) models, particularly gradient boosting and support vector machines (SVM), are better suited to model the complex, nonlinear relationships prevalent in large-scale health datasets.

Among individual models, the Pruned Gradient Boosting Machine (PGBM) achieved the highest sensitivity (90.8%) and a competitive AUC-ROC (0.94), outperforming both classical and baseline machine learning models. This supports previous findings by [2], [25], [41], who identified gradient boosting as one of the most accurate algorithms for predicting cardiovascular risk. Our enhancement of base learners—such as introducing pruning, regularization, and robust loss functions—further reduced overfitting and improved generalizability, aligning with best practices highlighted in [16] and [10].

The comparative advantage of ensemble models, particularly stacking, was striking.

The stacking ensemble achieved the best overall performance across all metrics (accuracy = 87.2%, sensitivity = 89.6%, specificity = 84.7%, AUC = 0.94). These results corroborate prior work by [22], [39], and [34], who emphasized the effectiveness of stacked models in reducing bias and variance through meta–learning. This can be illustrated by our hybrid stacking framework of combining interpretable models, e.g., logistic regression and more complex learners random forests and SVM, that shows how it is possible to maximize predictive performance and explainability simultaneously.

However, unlike in previous studies where the studies are mainly based on high-income populations [46], [8], our work relates to a significant literature gap by considering the model performance on a dataset that reflects risk profiles that are prevalent in LMICs. Even though it was based in the U.S., the use of the BRFSS dataset was justified methodologically because it covered behavioral and clinical variables (e.g., smoking, diabetes, hypertension) that are also common in LMICs (e.g., Kenya) [14], [27].

In addition, the adoption of newer ensemble methods, such as Bayesian Model Averaging (BMA) and majority voting, helped reduce the effect of class imbalance and made the model more stable in different demographic and clinical subgroups. This agrees with the findings of [33] [9] and [29] who inferred that the use of averaging-based ensembles could improve diagnostic performance among heterogeneous health populations.

Learning curve analysis and calibration curves were also used to give evidence of model reliability. Models like PGBM and HIRF were not only accurate but also had well-aligned predicted probabilities with observed cases, which is highly crucial in clinical practice where the determination of risks has to be highly reliable. The results of the study are consistent with those of the works by [18] and [42], which have found that well–calibrated interpretable AI models are essential to apply in practice in a real-life healthcare setting.

It is worth highlighting that the study also emphasizes the importance of interpretability tools, including the SHAP values, in explaining the model. Combined with ensemble predictions, SHAP explanations facilitated meaningful clinical interpretations, which justify using AI in settings where medical professionals need explicit reasons related to model decisions [4], [6].

This study has a few limitations that need to be considered despite the encouraging outcomes. First, although the BRFSS dataset provides a substantive size and a broad scope, the data provided is based on self-report, a phenomenon that can create potential bias in recall and poor clinical information, including those related to CHD or co-morbidities. Second, the data is US-based and might not completely represent regionally specific risk variables or allowance of healthcare accessions experienced in most LMICs. Although the selected features mirror common CHD risk indicators in LMICs, local validation using region-specific datasets is necessary to ensure model adaptability and contextual relevance. Additionally, some advanced ensemble techniques, such as stacking and Bayesian model averaging, require substantial computational resources and may pose implementation challenges in resource-constrained health systems without adequate infrastructure. Finally, while interpretability was improved using SHAP values and hybrid model design, black-box elements remain within some ensemble frameworks, potentially limiting clinician trust and adoption without further integration of explainable AI. Future research should focus on prospective validation, real-world deployment, and the use of locally collected datasets to enhance model transferability and clinical utility.

## Conclusion

This study presents a compelling case for the transformative potential of enhanced machine learning and ensemble models in the early prediction of CHD, particularly within the context of resource–limited healthcare systems. By systematically comparing classical statistical methods, baseline machine learning algorithms, and their enhanced counterparts–ultimately culminating in advanced hybrid ensemble strategies–our findings confirm that predictive accuracy, interpretability, and clinical utility are not mutually exclusive but can be achieved in tandem. The stacking ensemble, in particular, emerged as a standout performer, demonstrating not only superior classification metrics (AUC = 0.94; Sensitivity = 89.6%) but also operational adaptability through the integration of interpretable base models and explainability tools such as SHAP.

What distinguishes this study is its pragmatic design, which bridges the methodological rigor of machine learning with the pressing clinical needs of LMICs. Using the BRFSS dataset as a proxy for real-world health conditions and risk profiles prevalent in LMICs, we provide a scalable and transferable framework that can be localized with minimal resource demand. The implementation of synthetic oversampling, noise-resistant modeling, and model stacking collectively pushed the frontier of CHD prediction beyond conventional paradigms.

To support practical application, especially in low-resource clinical environments, the selected models were enhanced for interpretability and reduced computational overhead. Techniques such as SHAP values enable transparent decision–making, allowing clinicians to understand the rationale behind each prediction. This transparency fosters trust and supports clinical judgment. Moreover, lightweight variants (e.g., ANRDT and PGBM with early stopping) reduce training time and resource demands, facilitating deployment in environments with limited hardware or internet connectivity. Future work may focus on integrating these models into electronic health record systems or mobile health platforms for real-time screening and decision support.

We recommend that future research build upon this framework by deploying these models in real-world clinical decision-support systems (CDSS), integrating them with electronic health records (EHRs), and validating them using regional datasets from diverse healthcare environments. Although, the models were trained and validated on a large-scale dataset (n = 9,789), this study did not formally evaluate scalability metrics such as training time, inference latency, memory usage, or model size. Future research will incorporate benchmarking of computational efficiency and model complexity to assess real-time deployment feasibility in resource-constrained or low-latency environments.

## Data Availability

The data underlying the results presented in the study are available from IEEE DataPort (https://ieee-dataport.org/documents/heart-disease-dataset). The dataset is publicly accessible and was retrieved online. All data used in this study are secondary and were not collected by the authors. The data can be made available upon subscription from IEEE DataPort, or alternatively, provided by the authors upon reasonable request.

## Acknowledgments

The authors wish to acknowledge Titus Mtua Kioko and John Wafula Kiluyi for their valuable contributions in developing the Python code used to run the machine learning models in this study.

